# Evolution of Life-Threatening COVID-19 as the SARS-CoV-2 Pandemic Has Progressed

**DOI:** 10.1101/2023.02.15.23285759

**Authors:** Sudish C. Murthy, Ashley M. Lowry, Steven M. Gordon, Eugene H. Blackstone

## Abstract

**Rationale:** As the SARS-CoV-2 pandemic progressed, hospital mortality among patients admitted with COVID-19 decreased; however, its mechanism remains unclear.

**Objective:** To determine underlying factors that might explain the declining observed mortality of hospitalized patients with COVID-19.

**Methods:** This observational study used a prospective COVID-19 clinical database compiled by a 15-hospital health system. Participants were persons testing positive for SARS-CoV-2 (n=185,636), among whom 26,872 were hospitalized for COVID-19 from 3/1/2020 to 6/1/2022.

**Measurements:** Endpoints were hospital and 60-day mortality assessed by randomForests-for-survival machine learning using patient demographics, medical history, symptoms, and admission laboratory test results.

**Main Results:** Mortality of patients hospitalized with COVID-19 fell from 11% in 3/2020 to 3.7% in 3/2022; 60-day mortality was 17% in 5/2020 and 4.7% in 5/2022. Advanced age was the most important predictor of mortality, followed by admission laboratory test results. Risk-adjusted 60-day mortality was 15% had all patients been admitted in 3/2020, minimally unchanged at 12% had they been admitted in 5/2022. Dissociation between observed and predicted mortality was largely explained by change in admission patient profile, particularly admission laboratory test results. Transition to intensive care occurred rapidly for those predicted to do poorly.

**Conclusion:** Mortality from COVID-19 requiring hospitalization has declined as the pandemic has evolved, but surprisingly, persists for 60 days following admission. Demographics, medical history, and at-admission laboratory results continue to accurately predict mortality despite reduction of observed mortality and in spite of therapeutic advances. Importantly, rapid escalation of care can be predicted at admission using standardly obtained information. There has been a subtle but perceptible change in the at-risk population that explains these findings.

## INTRODUCTION

Few anticipated that severe acute respiratory syndrome coronavirus 2 (SARS-CoV-2)^1^ and its associated illness coronavirus disease 2019 (COVID-19)^2^ would still be a major public health problem 3 years after its clinical detection in December 2019.^3^ Identifying its genetic sequence synchronous with its clinical emergence,^4^ coupled with past experience with members of this viral family,^5^ led many to believe that the disease would soon pass. Unfortunately, despite rapid dissemination of information regarding the unique clinical characteristics and modes of transmission, early adoption of public health measures,^6^ and fairly rapid understanding of pathogenesis of severe disease,^7^ a pandemic emerged, only partially abated by rapid manufacture of vaccines.^8^

Nearly 100 million persons have contracted SARS-CoV-2 in the United States, with more than 1 million related deaths,^9^ thus it is important to understand changes in clinical profiles and outcomes as the pandemic has evolved given the emerging variants, widespread but incomplete population vaccination, and availability of antiviral therapy during the early stage of infection.^10^ Specifically, because of the observed reduction in hospital mortality from COVID-19, we questioned the effect of SARS-CoV-2 variant evolution, dissemination of vaccines, and development of advanced therapeutics as important inflection points in the pandemic. Finally, as patients were followed after hospital discharge, we were surprised by the number who had died soon after discharge, leading us to wonder if we knew the true magnitude of the illness in terms of associated mortality beyond hospitalization needed to help us prepare for what lies ahead in these still uncertain times.

Therefore, our aim with this study was to understand the risk of mortality of patients admitted to our health system hospitals with COVID-19 as the pandemic progressed by answering the following questions: 1) How did patients hospitalized with COVID-19 differ from non-hospitalized persons testing positive for SARS-CoV-2? 2) What was the mortality of patients in-hospital and early after hospital discharge? 3) What were the predictors of mortality? 4) Could the observed decrease in hospital mortality within our hospital system from COVID-19 over time be explained by the factors we studied and methodology we employed? We limited our investigation to what was known about these patients at hospital admission – their initial encounter – ignoring events and therapies in their subsequent hospital course, to help clinicians marshal appropriate resources at first contact with ill patients.

## PATIENTS AND METHODS

### Patients and Data

From 3/1/2020 to 6/1/2022, 185,636 people tested positive for SARS-CoV-2 in the Cleveland Clinic Health System. Of these, 26,872 (15%) were admitted with COVID-19 to 1 of 14 of its 15 Northeast Ohio and Florida hospitals (Table 1 and Figure 1). Early in the pandemic, our System established a COVID-19 Registry, capturing data from our electronic health records via automated feeds^11^ using standardized templates and manually by a study team. The Institutional Review Board (IRB) approved data collection with informed consent waived (IRB Record 136 under IRB 20-283, 4/27/2020).

**Table 1.**
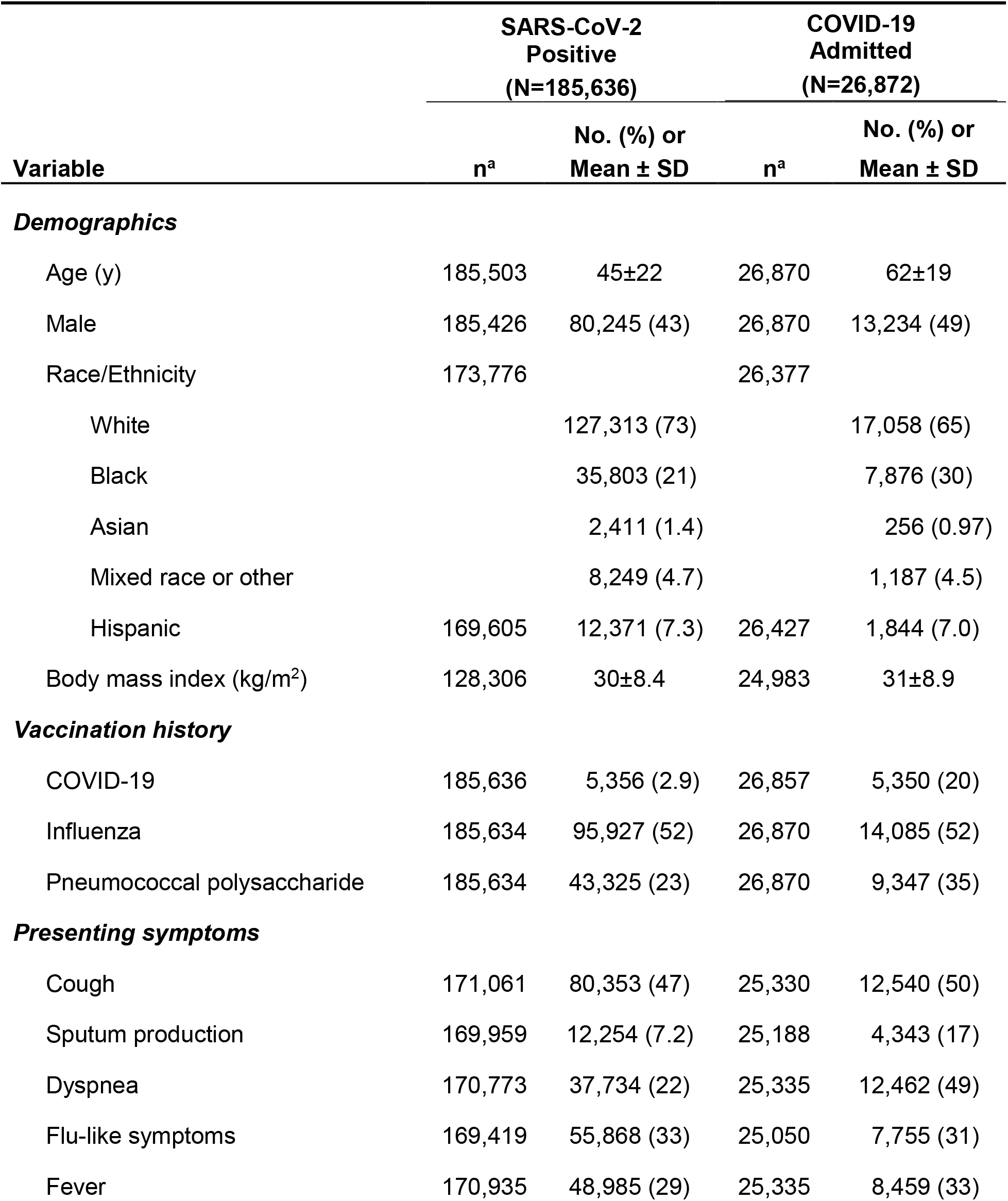
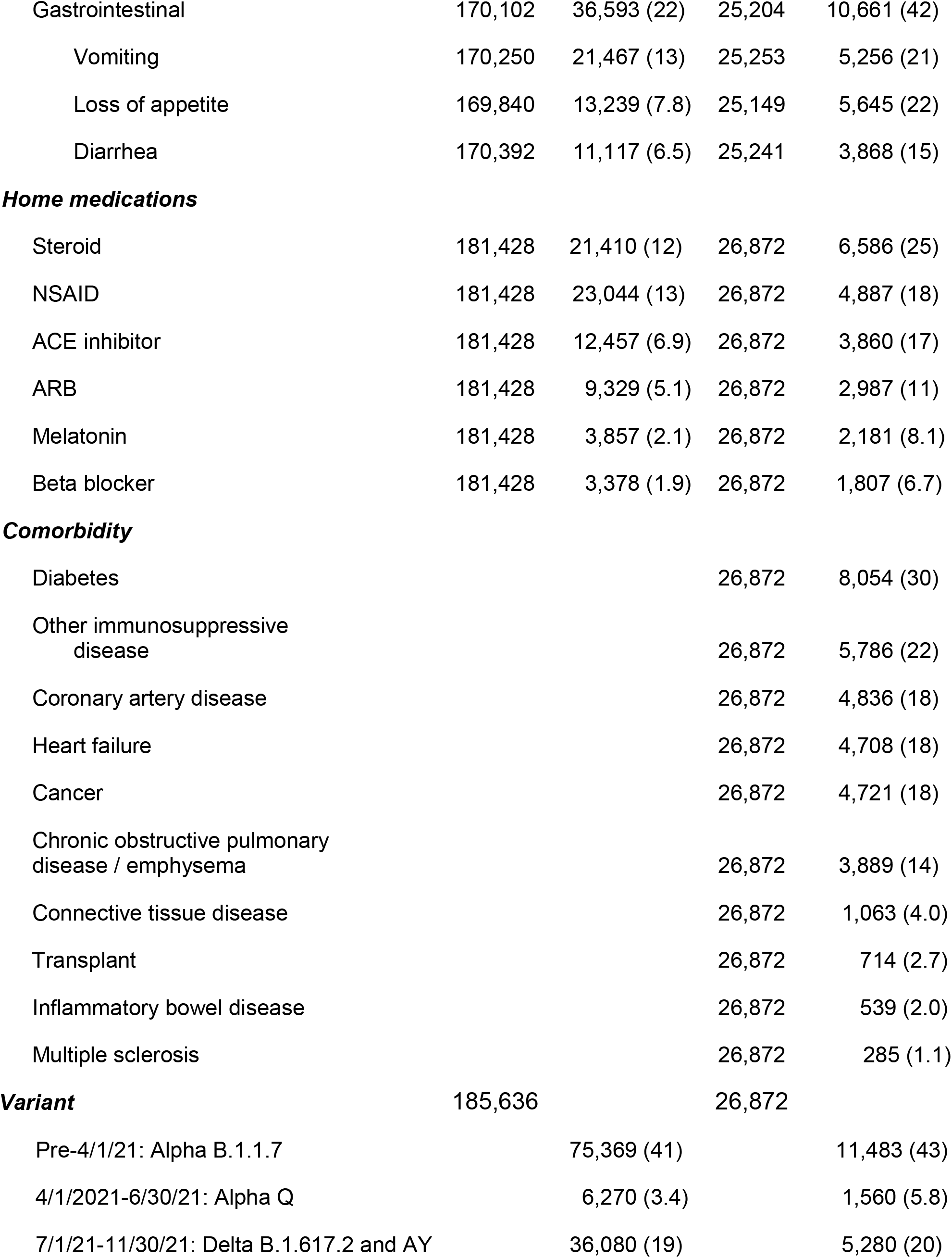
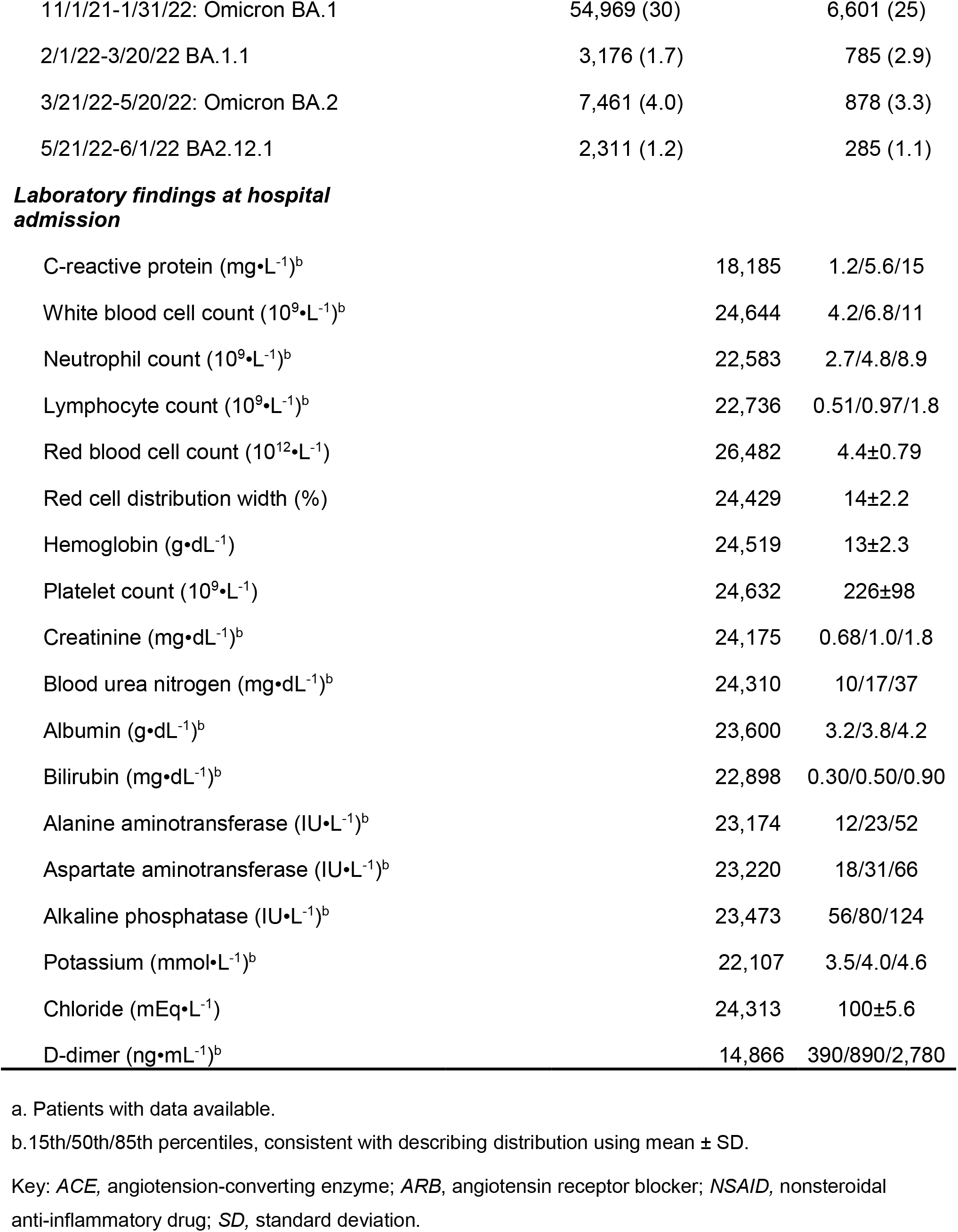
Characteristics and physiologic response to SARS-CoV-2 of persons testing positive and those admitted to Cleveland Clinic Health System hospitals with COVID-19

**Figure 1:**
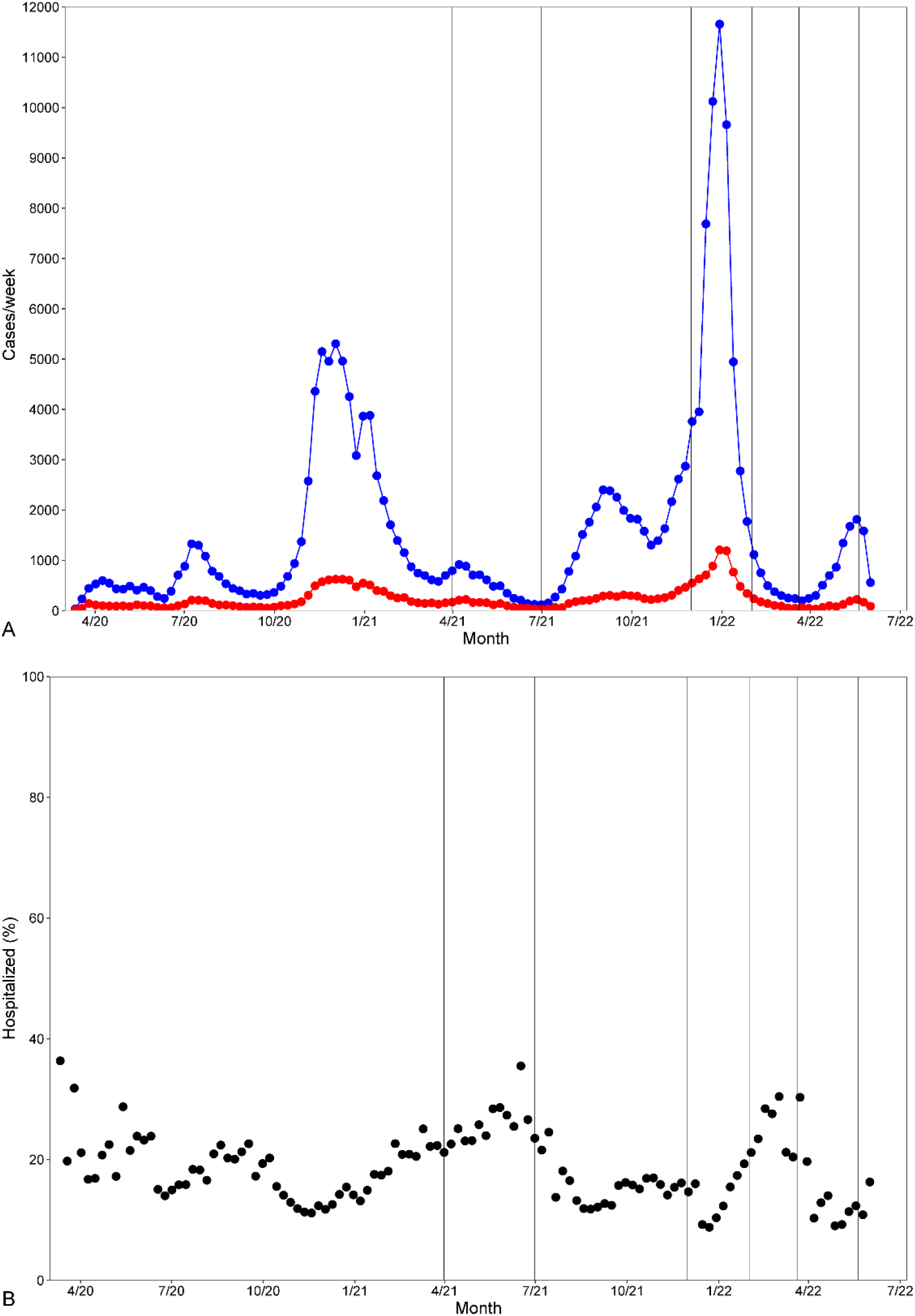
Trends in **A)** number of persons testing positive for SARS-CoV-2 from March 2020 to June 2022 (blue line and symbols) and number hospitalized for severe and life-threatening COVID-19 (red line and symbols), and **B)** percentage of persons testing positive for SARS-CoV-2 who were hospitalized. Each symbol represents a 1-week average. Vertical lines represent epochs of dominant SARS-CoV-2 variants:

- Pre-4/1/21: Alpha B.1.1.7
- 4/1/2021 to 7/1/21: Alpha Q
- 7/1/21 to 12/1/21: Delta B.1.617.2 and AY
- 12/1/21 to 2/1/22: Omicron BA.1
- 2/1/22 to 3/21/22: BA.1.1
- 3/21/22 to 5/21/22: Omicron BA.2
- 5/21/22 to 6/1/22: BA2.12.1

### Endpoints

The primary endpoint was all-cause mortality from date of hospital admission to 60 days or last date of follow-up if less than that for patients with incomplete follow-up. A secondary endpoint was admission to the intensive care unit (ICU). Discharged patients were followed at 2 weeks by a COVID home-monitoring program and subsequently by periodic telephone calls, use of MyChart (Epic Systems), and encounters recorded in the electronic medical record until January 1, 2022. Thereafter, Ohio patients were systematically followed for vital status using the state death registry, while Florida patients were less systematically followed. Yet 81% of patients discharged alive were followed at least 60 days after hospital admission (Figure E1). In-hospital events and discharge location are presented in Table E1.

### Data Analysis

SAS v9.4 (SAS Institute, Cary, NC) and R v4.0.0 (R Foundation for Statistical Computing, Vienna, Austria) were used for data analysis. Profile of SARS-CoV-2– positive persons and of COVID-19 patients at hospital admission are depicted by temporal trends across the pandemic. Time-related mortality after hospital admission was estimated parametrically by a temporal decomposition method to derive the instantaneous risk of death (hazard function).^12^

Two machine-learning analyses of time-related mortality were performed. The first, a “pre–COVID-19” model, incorporated demographics, medical history, medications, and vaccination status before SARS-CoV-2 infection (34 variables, Appendices 1 and 2); the second, an “at-admission” model, incorporated all the pre– COVID-19 variables plus symptoms, results of routine laboratory tests at hospital admission for COVID-19, and “variant-dominant” periods according to our Infectious Disease department (28 additional variables; Table 1, Appendices 1 and 2). For modeling, the R program randomForest-SRC was used, with missing data imputed on-the-fly.^13^ One thousand trees were grown with bootstrap samples of patients, and branch splits using a log-rank rule were based on a random subset of 6 or 8 variables, respectively, with a terminal node size of 15.

Predictive importance of variables was assessed by variable importance (VIMP), which hierarchically orders variables by magnitude of reduction of prediction error.^14^ Partial dependency plots were generated to visualize the relation of variables to outcome.^15^

To determine whether mortality of a patient admitted early in the pandemic differed from that had the same patient been admitted late in the series (virtual-twin analysis^16^), we incorporated date of hospital admission into random forest models. Sixty-day predicted mortality for each patient was then calculated as if that patient had been admitted on 3/1/2020 by substituting for actual date of admission this counterfactual date. This was repeated day-by-day to 6/1/2022 for the entire group of patients.

To investigate the difference between observed and predicted mortality, we performed a secondary analysis using all variables except date of admission. Then, probabilities of 60-day survival were predicted for each patient’s risk profile. These were aggregated into 7 risk categories to investigate change in risk profile across the pandemic and to characterize patients in each of these risk categories.

## RESULTS

### SARS-CoV-2–Positive and Hospitalized Individuals Across the Pandemic

People tested positive for SARS-CoV-2 in our Health System in waves, similar to the country as a whole, with multiple peaks and valleys and multiple viral variants (Figure 1A). The fraction of those hospitalized was 16% over the pandemic, but was higher early in the experience and again in the spring of 2021 and winter of 2022 (Figure 1B).

Average age of SARS-CoV-2–positive individuals decreased, then increased: from 65 years in 3/2020 to 55 in 5/2021 and 63 in 5/2022; average age of those hospitalized was 10 to 20 years older (Table 1 and Figure 2A). Early in the pandemic, more males than females were admitted, but this quickly approached 50:50 (Figure 2B), with slightly more females. Trends in race presented a complex pattern (Figures 2C and 2D), but with a higher proportion of Blacks hospitalized than non-hospitalized. More hospitalized patients than non-hospitalized ones were on medications for chronic diseases, commensurate with an older hospitalized population.

**Figure 2:**
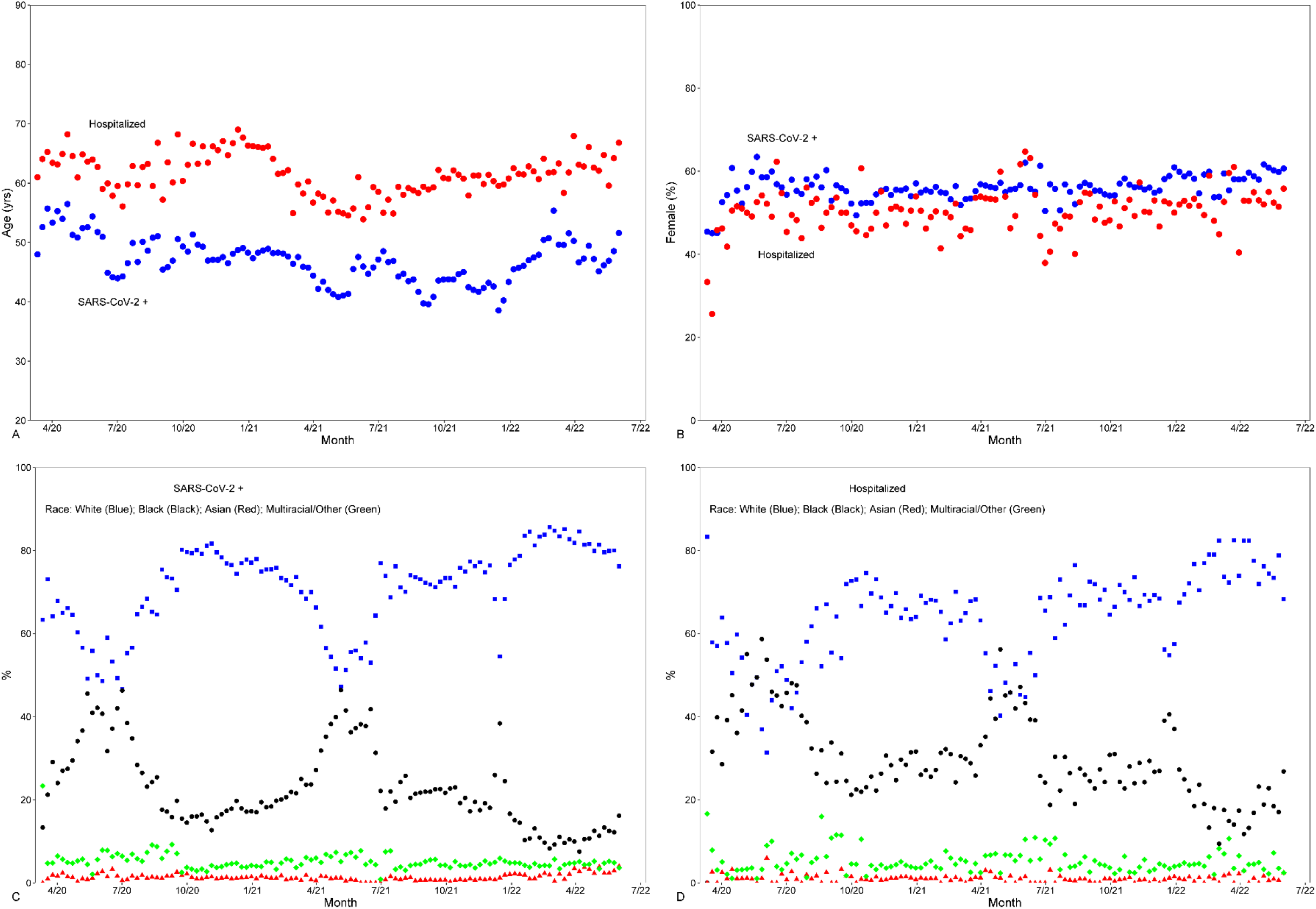
Trends in demographics. Symbols are 1-week averages. In panels *A* and *B*, blue symbols refer to persons testing positive for SARS-CoV-2 (SARS-CoV-2 +), and red symbols refer to patients admitted with COVID-19. **A)** Mean age. **B)** Proportion female. **C)** Race of persons testing positive for SARS-CoV-2. Key: White (Blue); Black (Black); Asian (Red); Multiracial/Other (Green). **D)** Race of persons hospitalized with COVID-19 (color key as in panel *C*).

Symptoms among both SARS-CoV-2–positive and hospitalized patients changed substantially across the pandemic, with self-reported fever and cough progressively declining (Figures 3A and 3B). Hospitalized patients were more symptomatic than non-hospitalized individuals, particularly with dyspnea and gastrointestinal symptoms (Figures 3C and 3D).

**Figure 3:**
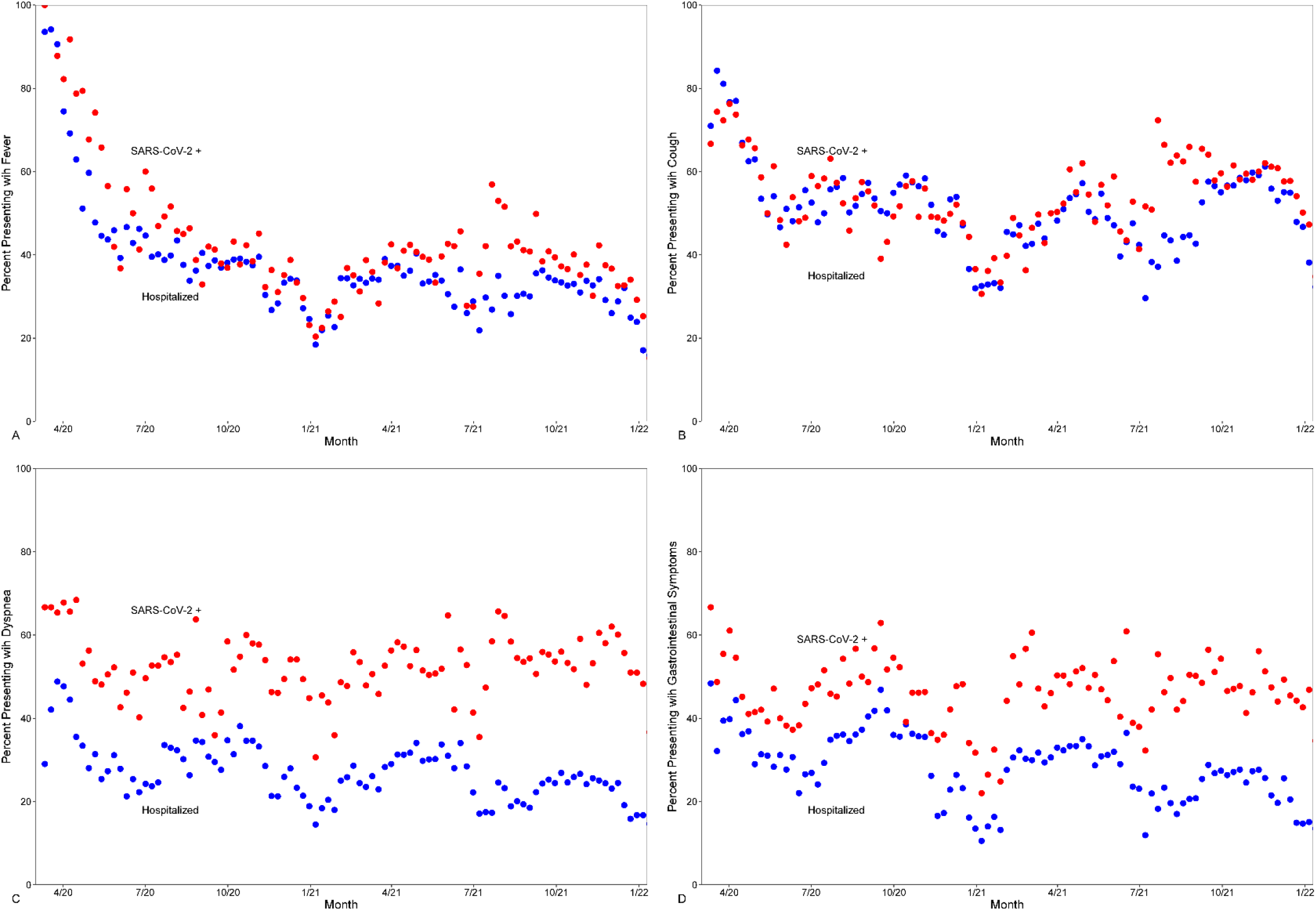
Trends in symptoms of persons testing positive for SARS-CoV-2 (SARS-CoV-2 +, blue symbols) and of patients hospitalized for severe or life-threatening COVID-19 (red symbols). Symbols are 1-week averages. **A)** Fever. **B)** Cough. **C)** Dyspnea. **D)** Gastrointestinal symptoms. Format as in Figure 2, panels *A* and *B*.

Results of routine laboratory tests at hospital admission also fluctuated across the pandemic, somewhat synchronous with disease surges (Figure 4). In particular, C-reactive protein declined early in the experience and more so in patients admitted in 2022 (Figure 4D), as did hepatic function enzymes (Figure 4E).

**Figure 4:**
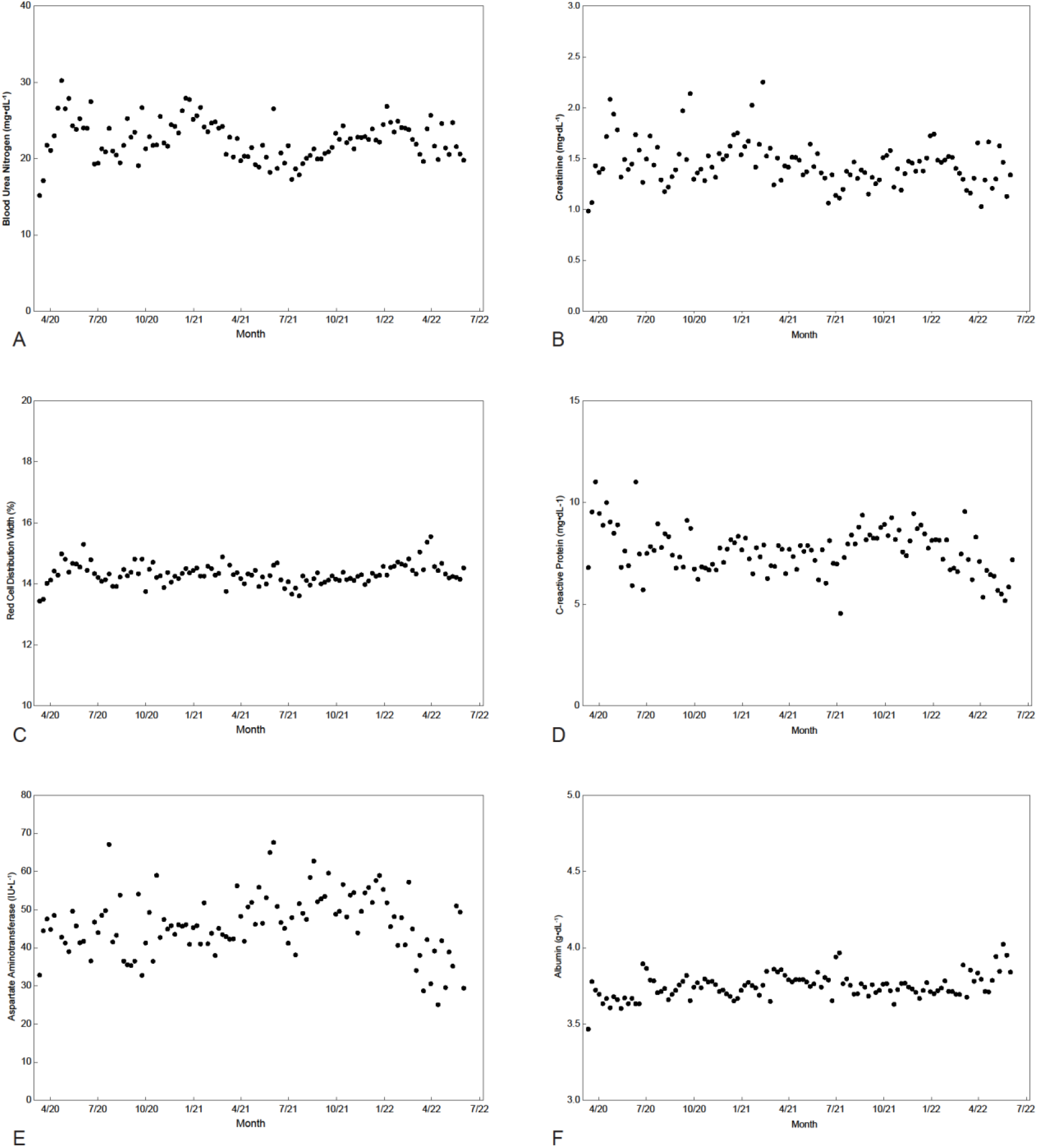
Temporal trends in physiologic response to SARS-CoV-2 at hospital admission for COVID-19. Symbols are 1-week averages. **A)** Blood urea nitrogen. **B)** Creatinine. **C)** Red cell distribution width. **D)** C-reactive protein. **E)** Aspartate aminotransferase. **F)** Albumin.

### Mortality Across the Pandemic

Observed hospital mortality fell from 11% in 3/2020 to 7.2% in 9/2020, 4.8% in 3/2021, 4.9% in 9/2021, and 3.7% in 3/2022 (Figure 5). However, mortality continued after hospital discharge: 9.9% at 30 days, 12% at 60 days, 13% at 90 days, 15% at 180 days, and 17% at 360 days (Figure 6). Instantaneous (unadjusted) risk of death peaked at 0.45%/day 9 days after hospital admission (Figure 6, inset), falling thereafter to reach 0.037%/day at 60 days, 4.0 times greater than an age-sex-race–matched U.S. population reference of 0.0093%/day. Actuarial mortality was highest for alpha variant B.1.1.7, lower for delta B.1 and omicron BA.1, and lowest for alpha-Q B.1.1 and omicron BA.2 (Figure 7A), although after risk adjustment, these differences were small (Figure 7B).

**Figure 5:**
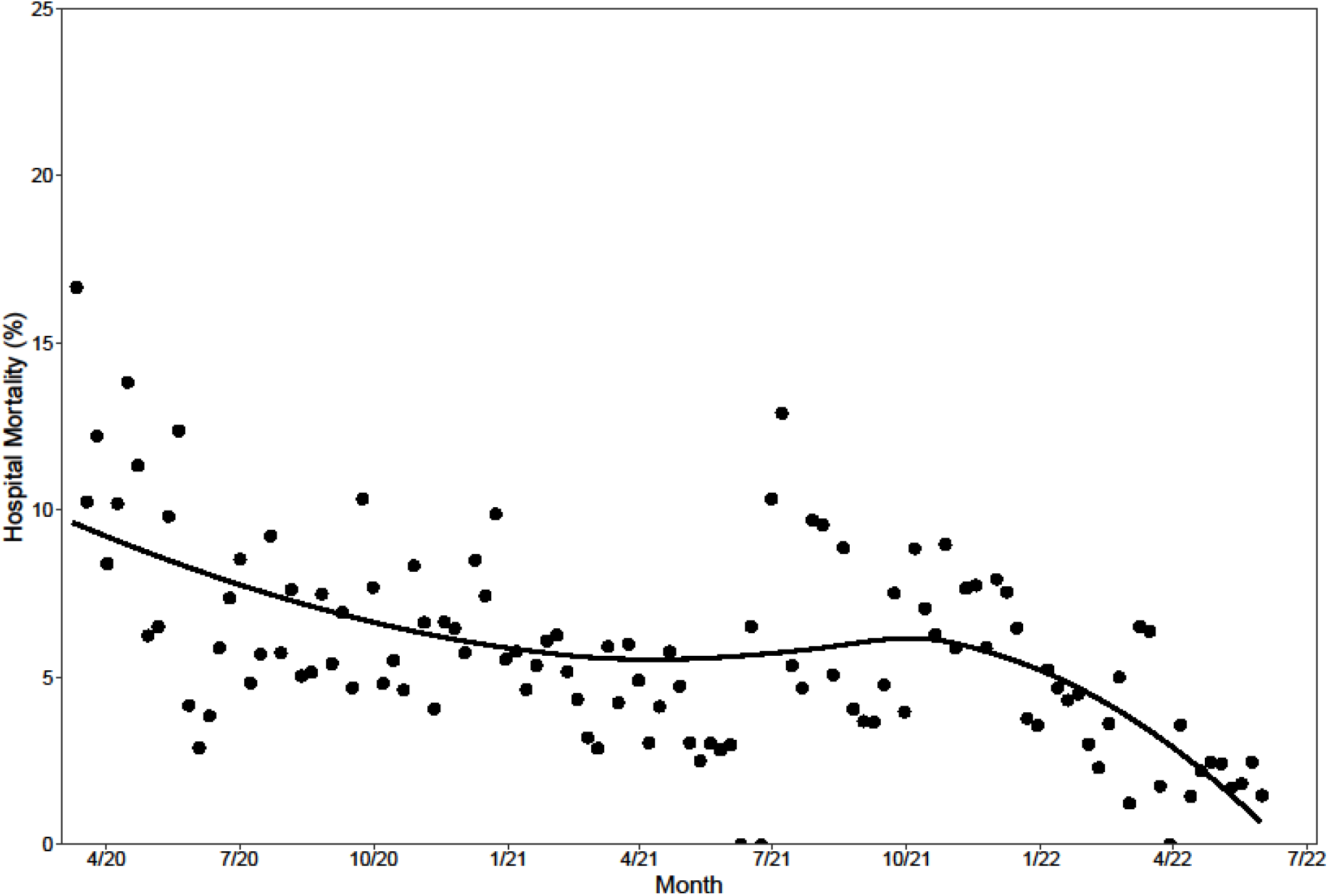
Weekly hospital mortality of patients admitted with COVID-19. Symbols depict actual mortality, and solid line is a loess (locally weighted scatterplot) smoother.

**Figure 6:**
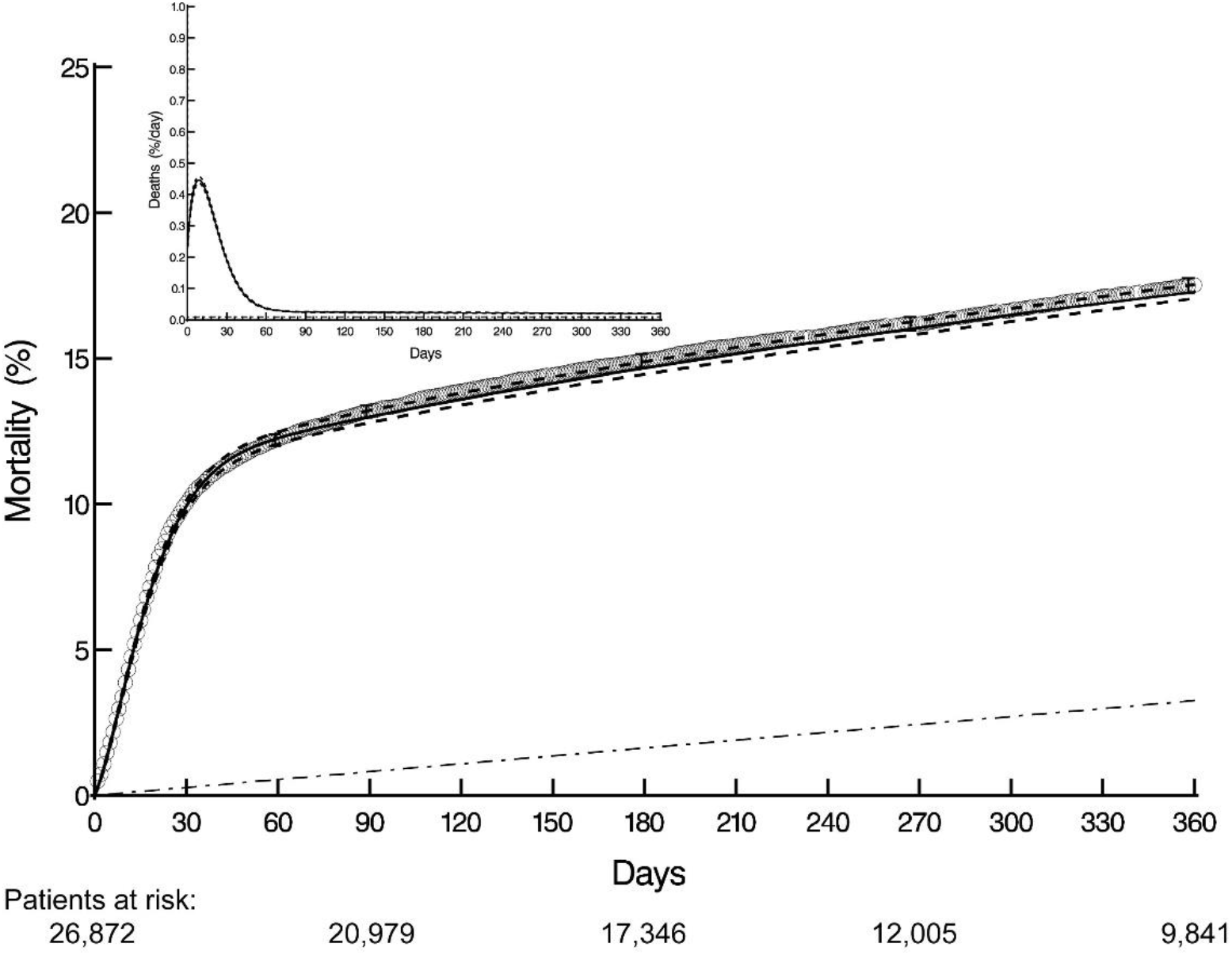
Time-related mortality of patients hospitalized with COVID-19. Each circle represents a death; vertical bars are 68% confidence limits (equivalent to ±1 standard error), and solid line enclosed within a 68% confidence band represents parametric estimates from which the instantaneous risk of mortality (inset) was derived. Number of patients remaining at risk at various intervals is shown beneath horizontal axis. Dash-dot-dash line represents mortality of an age-sex-race–matched U.S. population. Note that the early hazard extends out to at least 60 days.

**Figure 7.**
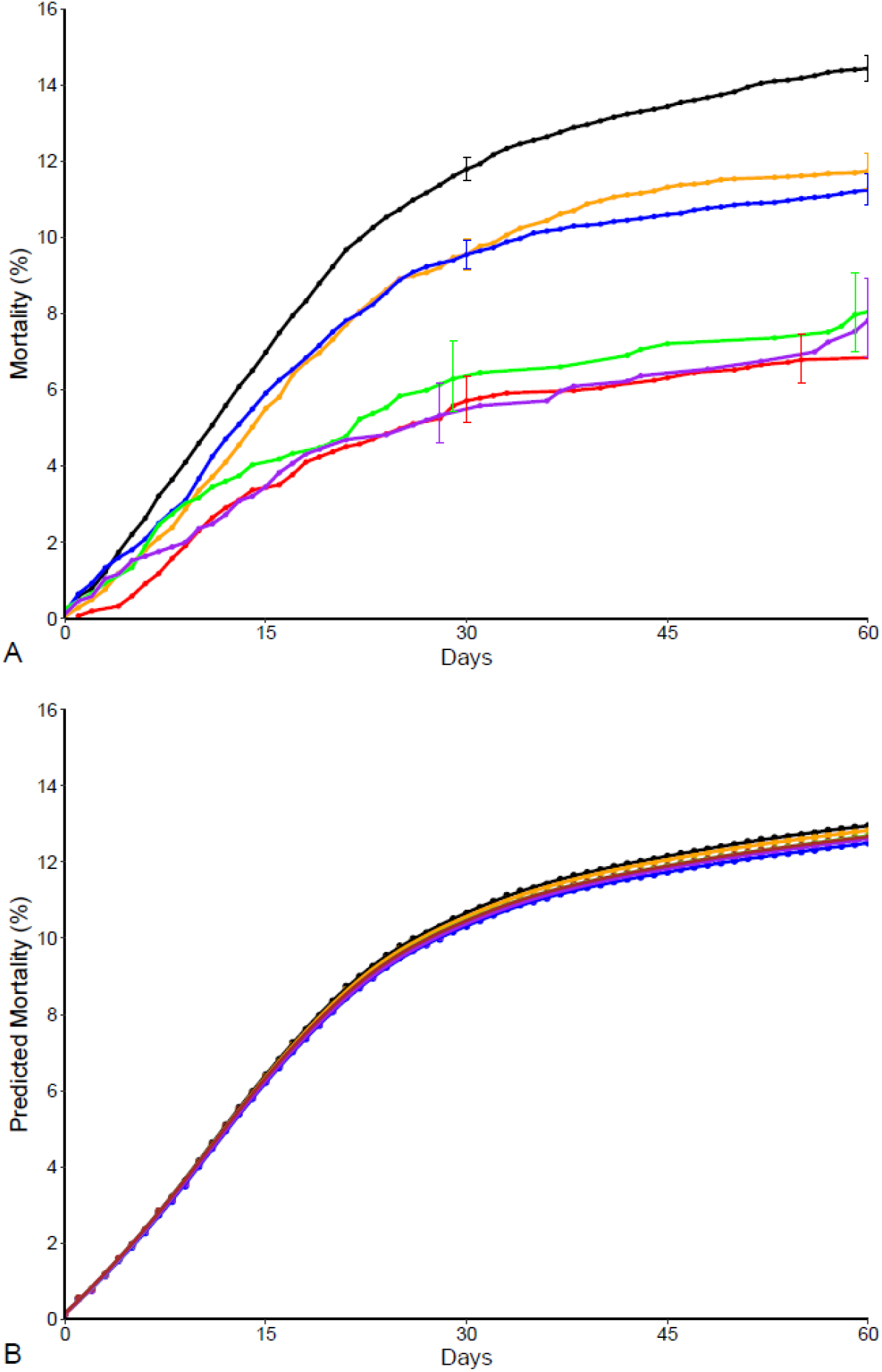
Mortality after hospital admission for COVID-19 according to SARS-CoV-2 variant. **A)** Unadjusted Kaplan-Meier estimates with 68% confidence limits (bars) equivalent to ±1 standard error, representing each variant (see key). **B)** Partial dependency plot of mortality according to SARS-CoV-2 variant from at-admission model. **Key:**

- Black: Pre-4/1/21: Alpha B.1.1.7; n = 11,483 (43%)
- Red: 4/1/2021 to 7/1/21: Alpha Q; n = 1,560 (5.8%)
- Orange: 7/1/21 to 12/1/21: Delta B.1.617.2 and AY; n = 5,280 (20%)
- Blue: 12/1/21 to 2/1/22: Omicron BA.1; n = 6,601 (25%)
- Green: 2/1/22 to 3/21/22: BA.1.1; n = 785 (2.9%)
- Purple: 3/21/22 to 5/21/22: Omicron BA.2; n = 878 (3.3%)
- Brown: 5/21/22-6/1/22: BA2.12.1; n = 285 (1.1%)

### Predictors of 60-Day Mortality

#### Pre–COVID-19 Model

Older age was the most important predictor of mortality (Figures 8A and 8B), with an upward inflection beginning about age 50 years, followed by a rapid increase with each year of age (Figure E2). It eclipsed the importance of body size, race, and heart and chronic pulmonary diseases. Neither SARS-CoV-2 vaccination nor SARS-CoV-2 variant was importantly associated with mortality (Figures 8A, 8C, and 9, the latter showing the small fraction of hospitalized patients who had been vaccinated at least 2 weeks prior to hospital admission).

**Figure 8:**
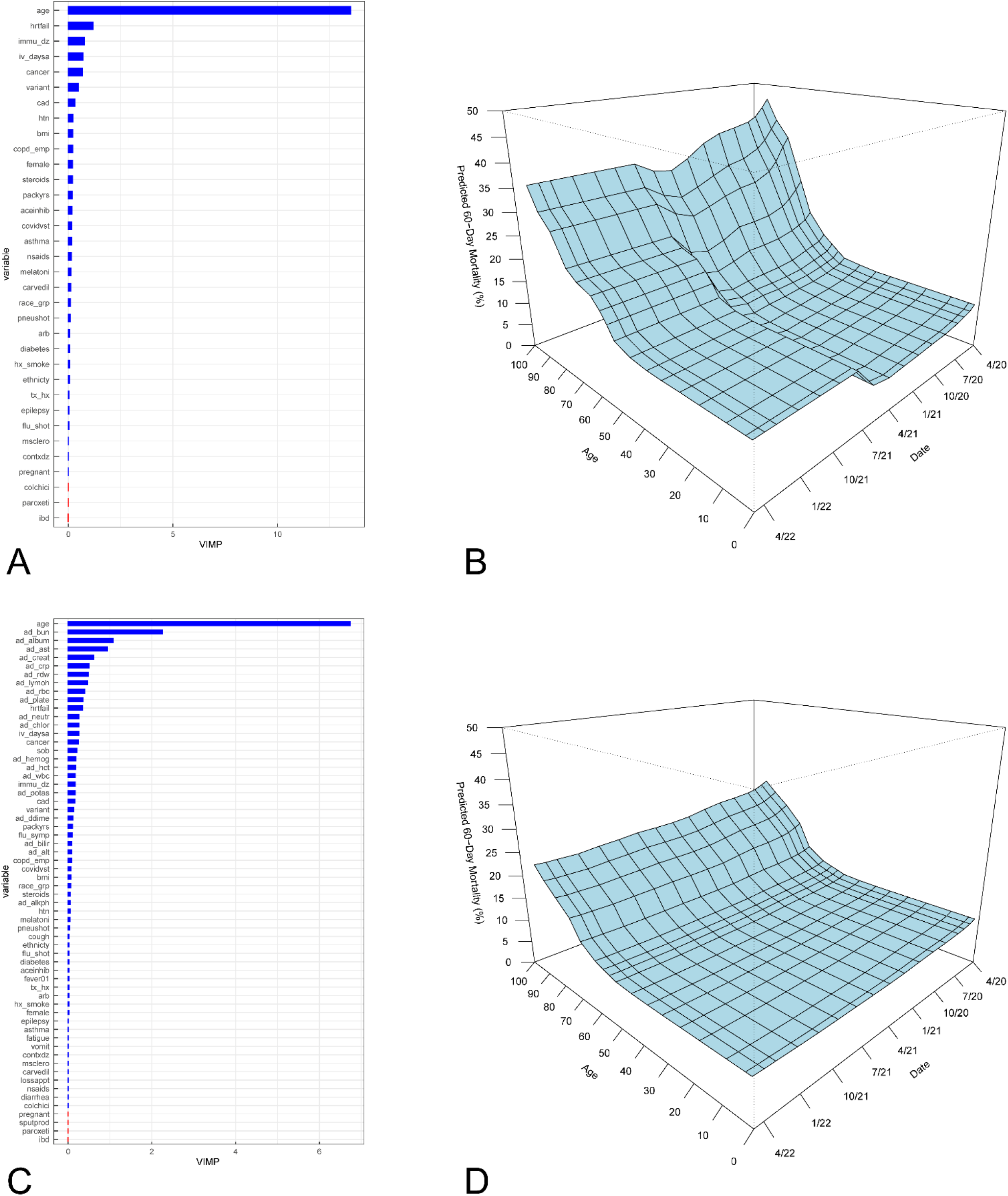
Variable importance (VIMP) for models of mortality. **A)** VIMP for pre–COVID-19 model of mortality (error=0.25). Key to variable names along vertical axis is provided in Appendices 1 and 2. **B)** Partial dependence plot of predicted 60-day mortality with respect to both age and date of hospital admission, based on pre–COVID-19 random forest analysis shown in panel *A*. “Days” refers to hospital admission date expressed as number of days since March 1, 2020. **C)** VIMP for at-admission model of mortality (error=0.18). Notice that after age, the most important variables are admission laboratory values reflecting physiologic response to SARS-CoV-2. **D)** Partial dependence plot of predicted 60-day mortality with respect to both age and date of hospital admission, based on at-admission random forest analysis shown in panel *C*.

**Figure 9:**
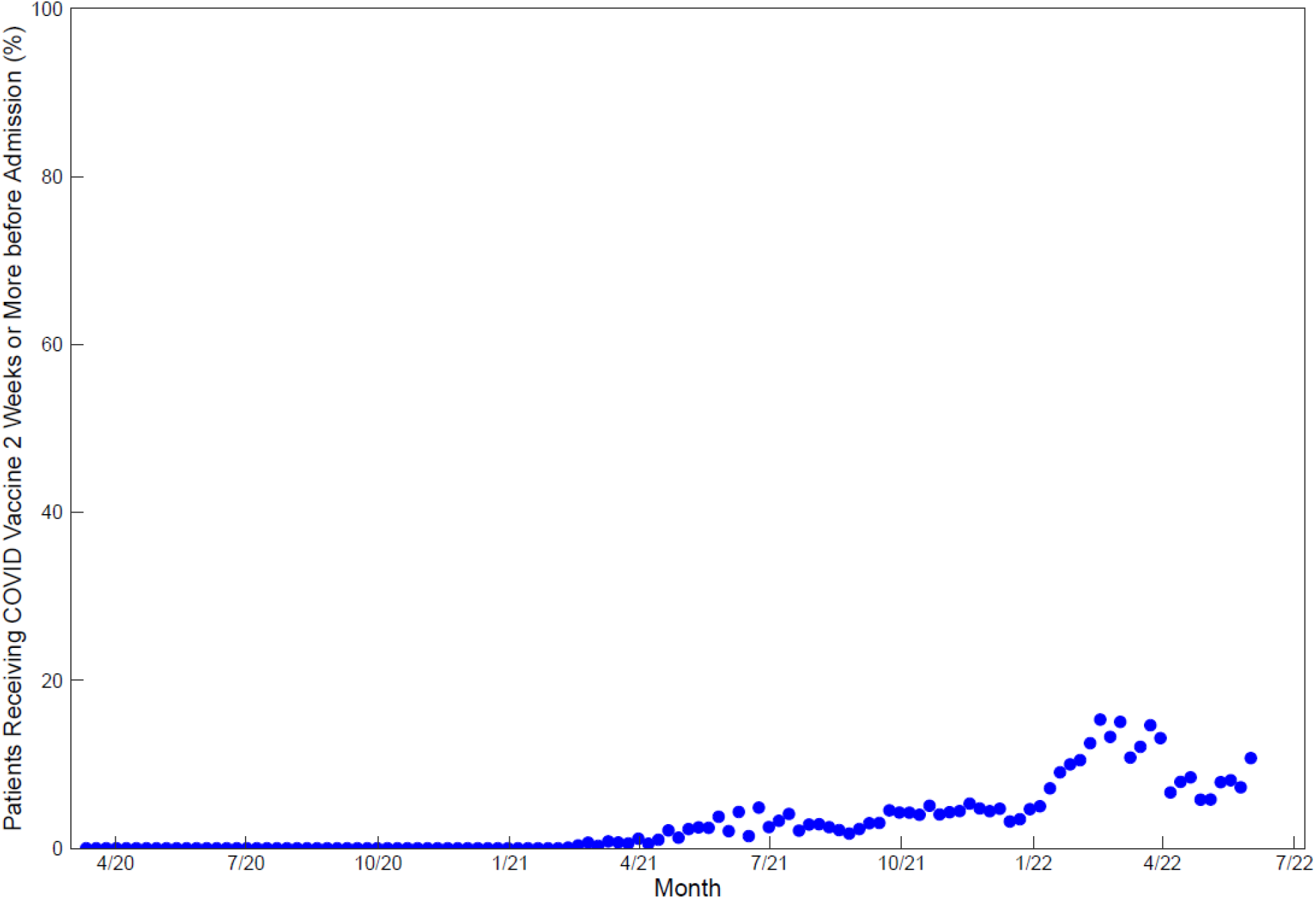
Percent of hospitalized patients vaccinated against SARS-CoV-2. Note that persons vaccinated within 2 weeks of hospital admission with COVID-19 were not considered fully vaccinated for this analysis.

#### At-Admission Model

When the presenting symptoms and results of routine-admission laboratory tests were added to the pre–COVID-19 model (Figures 8C and 8D), older age remained the most important predictor of mortality (Figure E2), but its importance was somewhat blunted (Figures 2B, 2D, and E2). Rather, several laboratory values at hospital admission emerged as important risk factors. Higher values of renal and liver dysfunction, greater systemic inflammatory response, abnormal hematologic values, and low albumin levels were associated with higher mortality (Figure 10).

**Figure 10:**
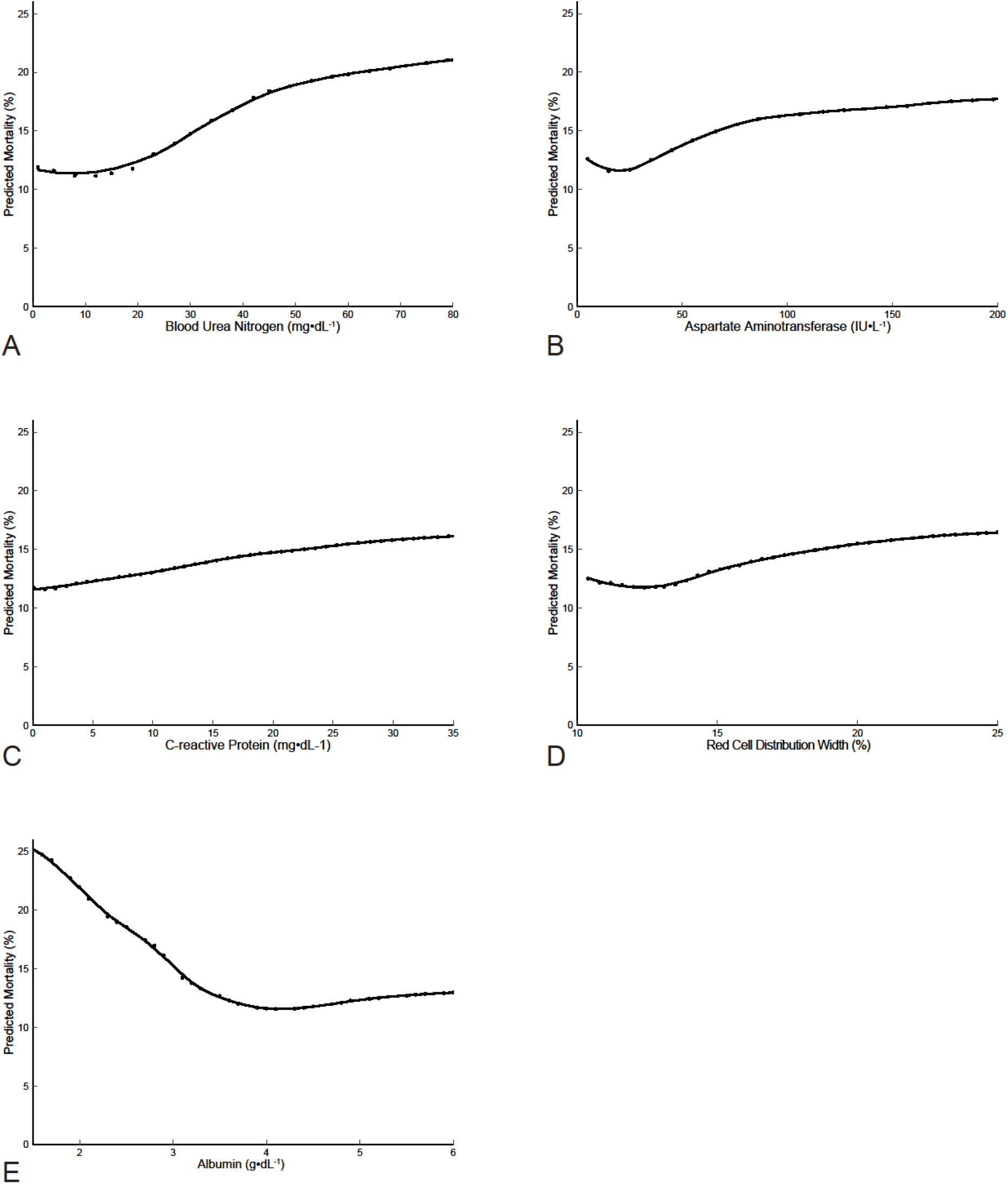
Partial dependency plots of predicted 60-day mortality according to routine laboratory findings at hospital admission for COVID-19. **A)** Blood urea nitrogen. **B)** Aspartate transaminase. **C)** C-reactive protein. **D)** Red blood cell distribution width. **E)** Albumin.

### Unchanging Risk Factors, But Changing Patient Profile, Lead to Changes in Mortality

Based on the at-admission model, 60-day predicted mortality from date of admission had all 26,872 patients been admitted in 3/2020 was 15%, in 5/2020 14%, in 11/2020 13%, in 5/2021 12%, in 11/2021 12%, and in 5/2022 12%, essentially unchanged over the pandemic (Figure 11). These small changes were far less than the observed actuarial trajectory of mortality: 17% in 5/2020, 14% in 11/2020, 5.2% in 5/2021, 12% in 11/2021, and 4.7% in 5/2022. Divergence of observed and risk-adjusted predicted mortality—a Simpson’s Paradox^17^—was attributable both to changes across the pandemic in the profile of demographics and pre–COVID-19 conditions, but in particular to changes in the profile of patients’ physiologic response to the disease as assessed by laboratory values at hospital admission (Figure 12A). Specifically, when coalesced into 7 predicted risk groups, progressively fewer highest risk patients were admitted over time, which contributed to the decline in observed mortality. Simultaneously, there was a mild reduction of the lowest risk patients, but expansion of patients with intermediate risk profiles, narrowing the risk profile of admitted patients across the pandemic.

**Figure 11.**
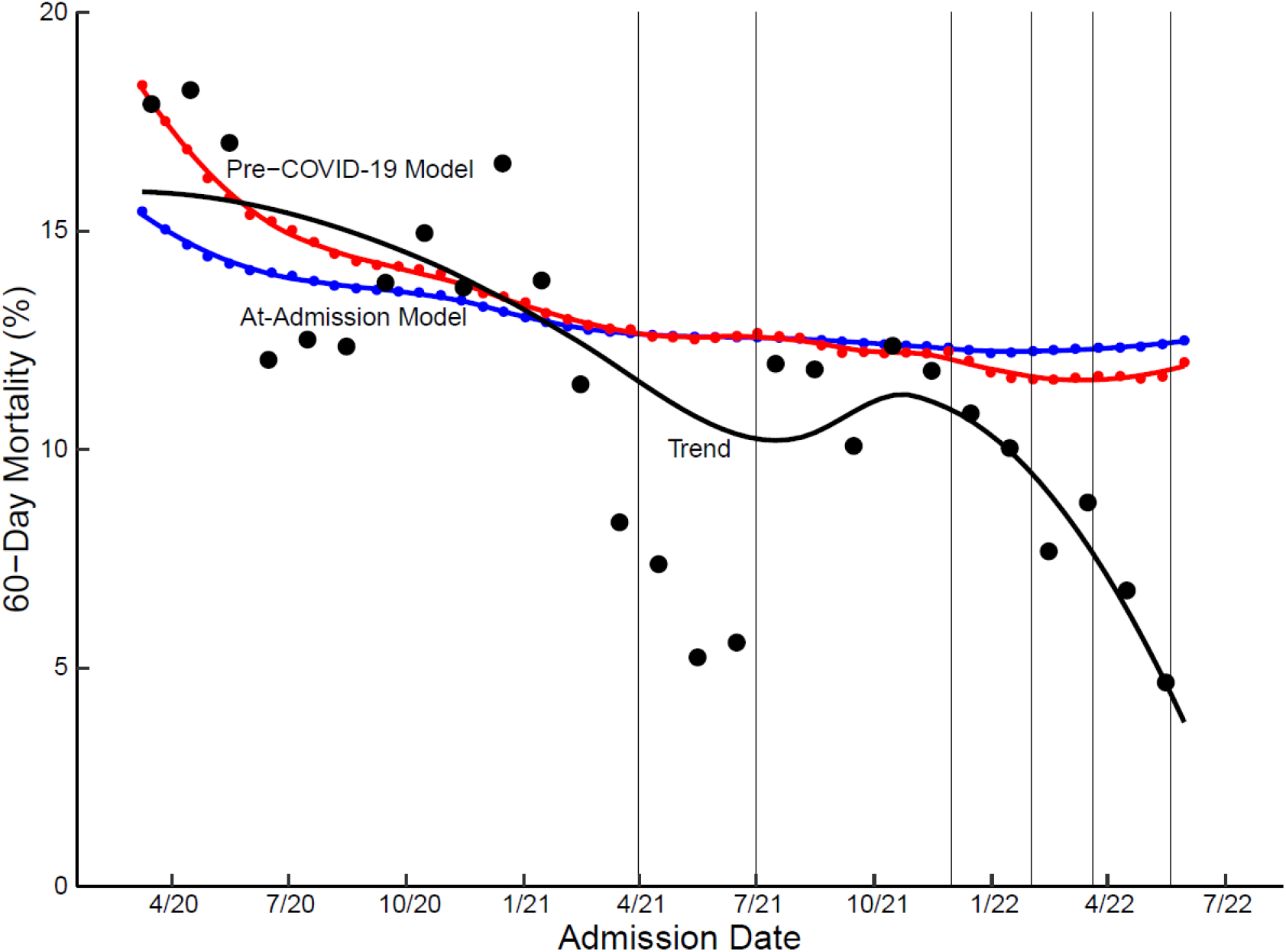
Actuarial and simulated 60-day mortality of patients hospitalized with COVID-19 across the pandemic. Black symbols represent observed 60-day unadjusted actuarial mortality for patients admitted each month, and black line is a loess (locally weighted scatterplot) smoother. Red and blue curves represent simulated 60-day mortality if all hospitalized patients across the pandemic were admitted on the same date from 3/1/2020 to 6/1/2022, according to pre–COVID-19 (red curve) and at-admission (blue curve) mortality models. Vertical lines denote approximate epochs of SARS-CoV-2 variants.

- Pre-4/1/21: Alpha B.1.1.7
- 4/1/2021 to 7/1/21: Alpha Q
- 7/1/21 to 12/1/21: Delta B.1.617.2 and AY
- 12/1/21 to 2/1/22: Omicron BA.1
- 2/1/22 to 3/21/22: BA.1.1
- 3/21/22 to 5/21/22: Omicron BA.2
- 5/21/22 to 6/1/22: BA2.12.1

**Figure 12:**
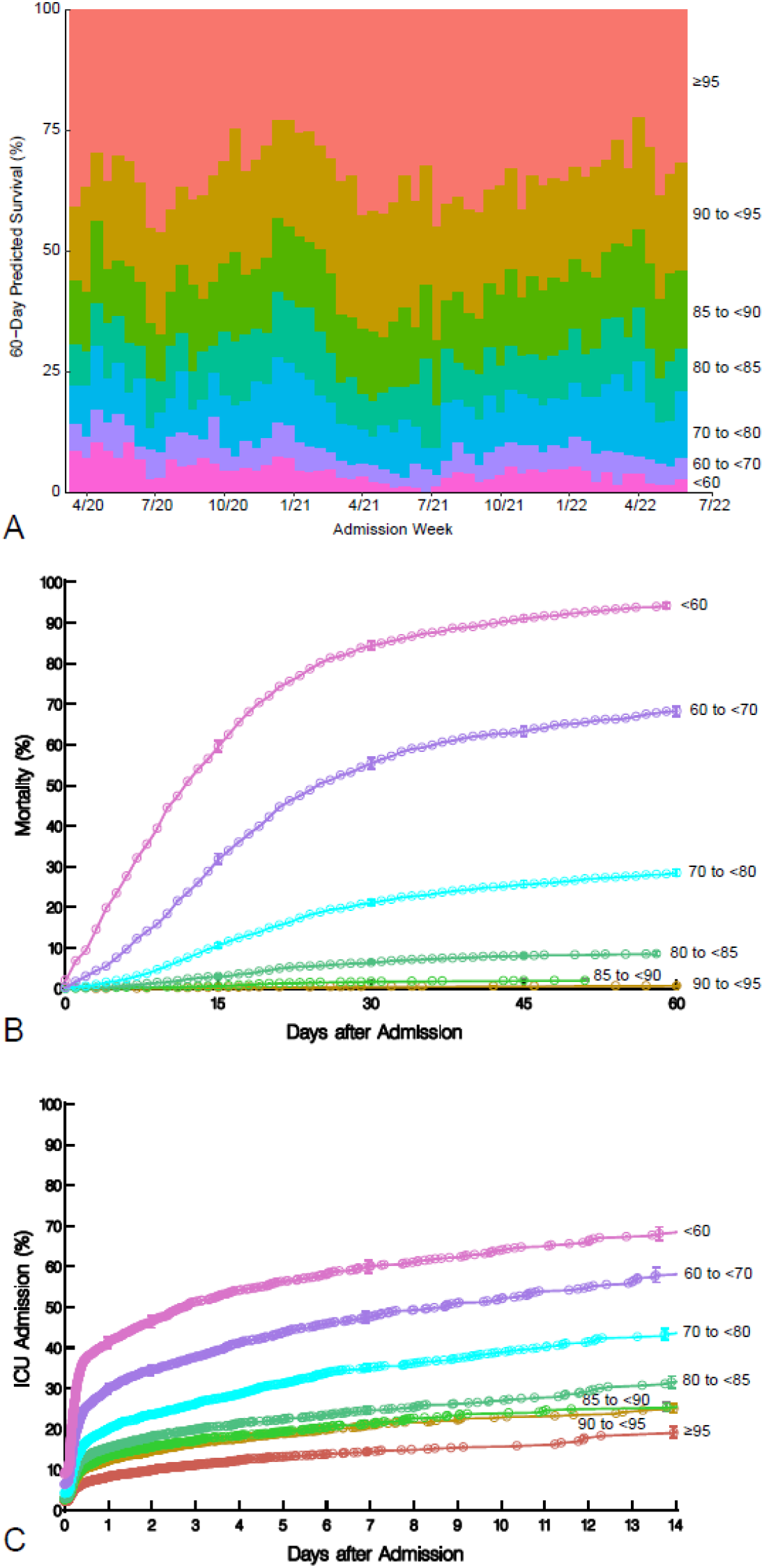
Distribution of 7 risk profiles of patients with COVID-19 across date of hospital admission and observed survival for each profile. **A**, Predicted 60-day survival using the at-admission model for mortality for profiles 1-7 are as follows: (1) ≥95%, (2) 90% to <95%, (3) 85% to <90%, (4) 80% to <85%, (5) 70% to <80%, (6) 60% to <70%, (7) <60%. Note particularly the gradual reduction in percentage of hospital admissions of the highest risk patients (pink and purple) and increase in number of patients in lower risk categories. **B,** Observed mortality stratified by predicted risk profile. **C,** Observed escalation of care to intensive care unit (ICU). In panels *B* and *C*, each symbol represents an event estimated by the Kaplan-Meier method, and vertical bars are 68% confidence intervals equivalent to ±1 standard error.

Actuarial survival of patients in these 7 risk groups demonstrated a large separation, particularly among the 3 highest risk groups (predicted survival less than 80%) (Figure 12B). The highest risk groups were elderly patients, more often White, with many comorbidities being treated medically, higher C-reactive protein, and elevated neutrophil counts but lower lymphocyte counts (Table 2). These same patients had the highest likelihood of being escalated to intensive care within 24 hours of admission (Figure 12C). In contrast, the lowest risk group (34% of hospitalized patients) had a median age 48, had few comorbidities and were taking few medications, and had few derangements of their at-admission laboratory profile.

**Table 2.**
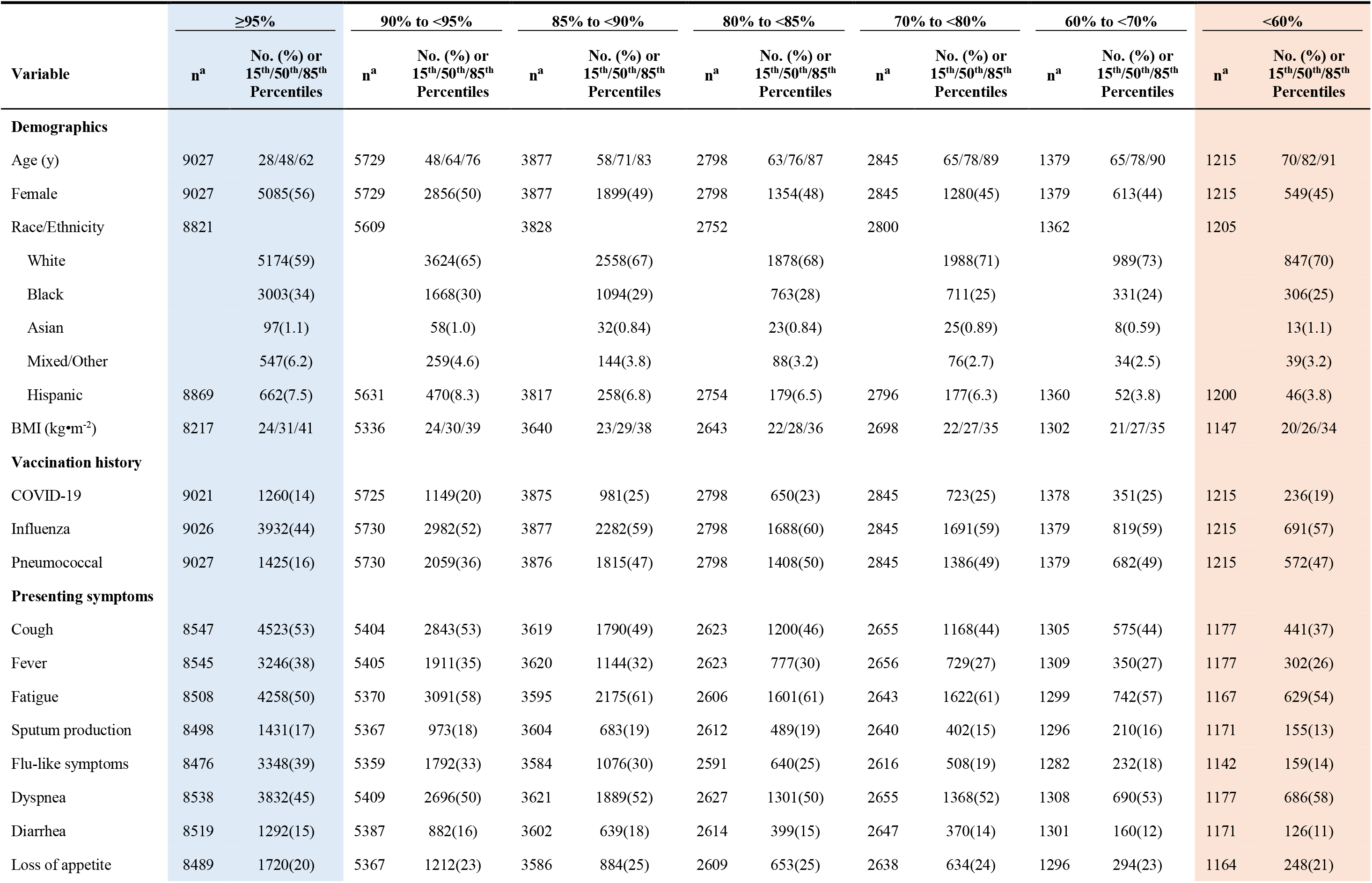
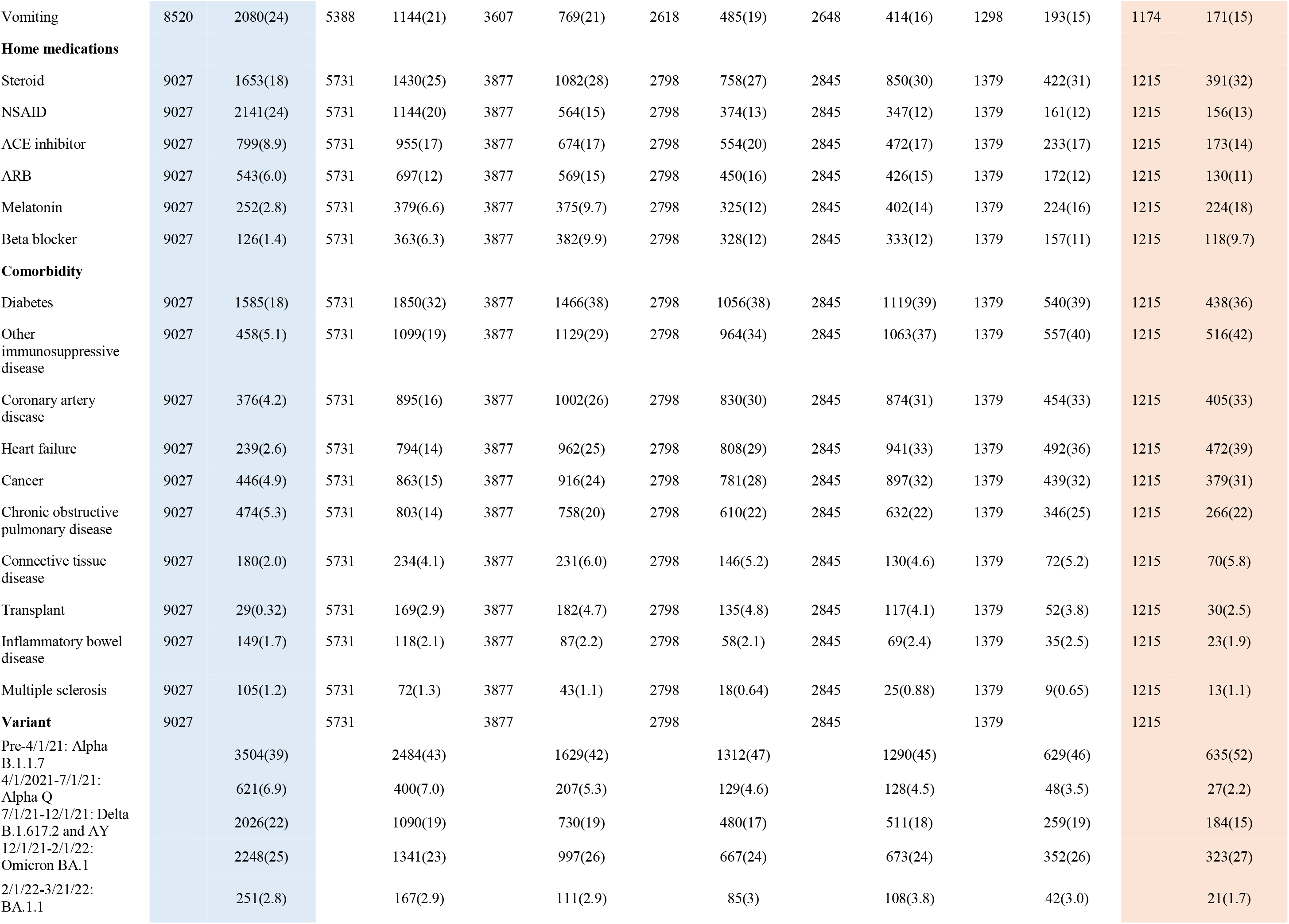
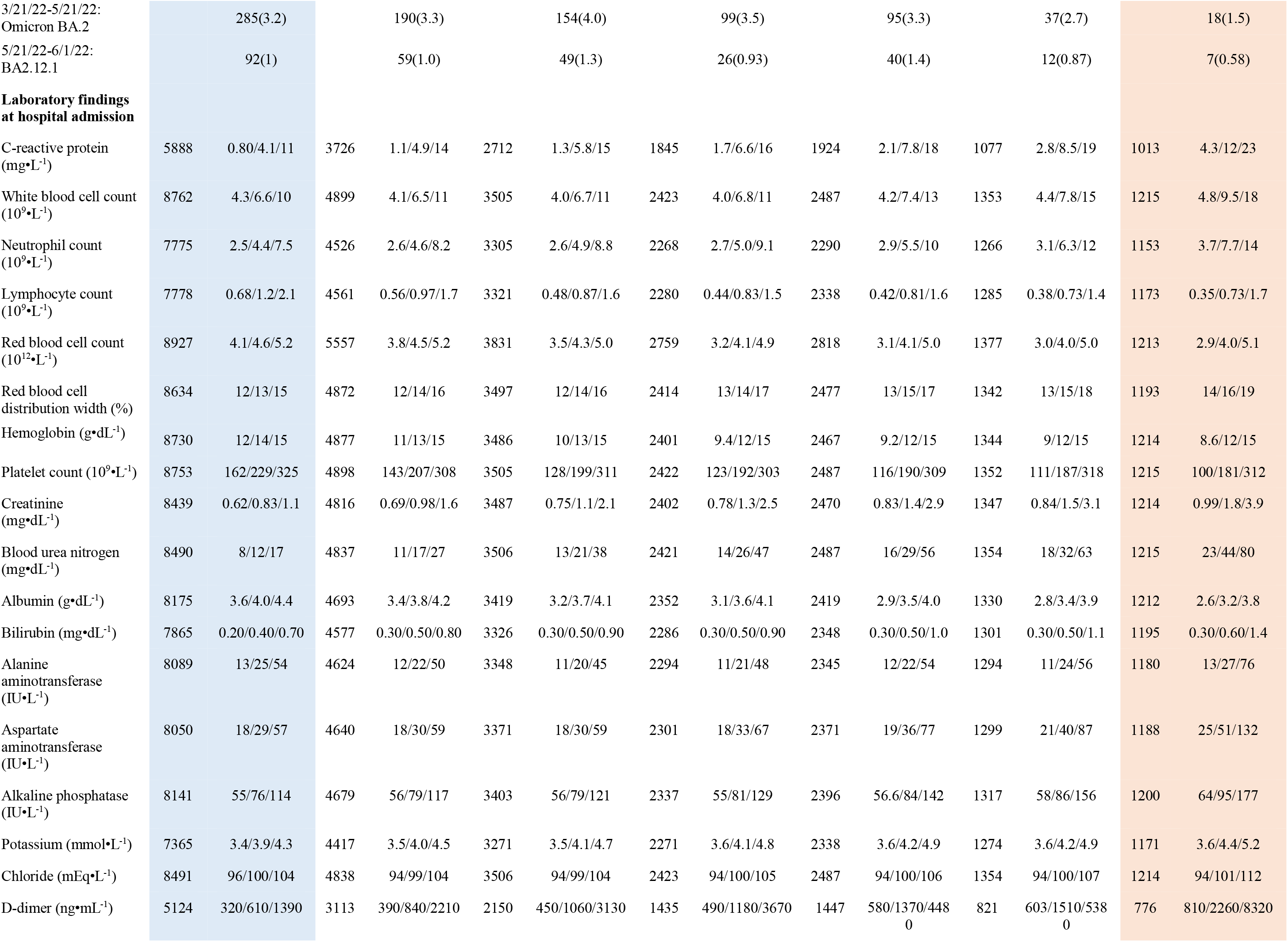
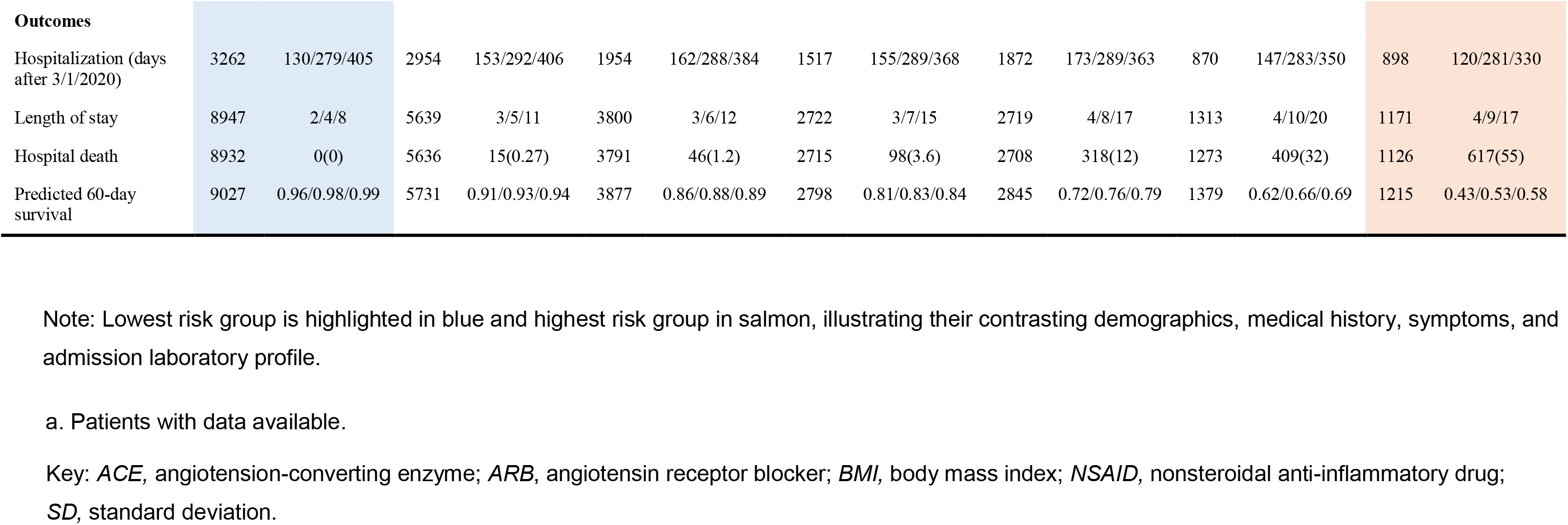
Profile of hospitalized patients according to 7 predicted 60-day survival categories by at-admission model: (1) ≥95%, (2) 90% to <95%, (3) 85% to <90%, (4) 80% to <85%, (5) 70% to <80%, (6) 60% to <70%, and (7) <60% (see Figure 11, A). Blue and salmon shaded columns highlight contrasting patient characteristics between lowest and highest risk groups.

## DISCUSSION

Undoubtedly, mortality among patients hospitalized with COVD-19 is trending downward. Our longitudinal assessment of these patients, both singularly and as a population, has allowed us to gain unique perspectives as this pandemic has evolved. We have been able to predict rapid deterioration of certain hospitalized patients based on a single assessment performed at hospital admission and to discover a continued post-hospitalization mortality from the disease. In addition, we have gained some clarity on the effectiveness of public health efforts and how hospital-requiring COVID-19 has changed as vaccination, preadmission therapeutics, and variants have permeated the landscape. Moreover, we now have insight into what to expect in the future as this pandemic morphs into an endemic disease.

### Principal Findings

There is decreasing observed 60-day mortality among patients hospitalized with COVID-19. Despite this, mortality for every risk profile based on routine admission assessment has decreased minimally. Thus, likelihood of mortality remains eminently predictable at hospital admission. Prediction accuracy is substantially refined by early physiologic response, as measured by routine admission laboratory tests, in addition to patients’ demographics and preexisting medical conditions, despite continued evolution of SARS-CoV-2 variants and an increasingly vaccinated population. Appearance of new SARS-CoV-2 variants, advancements in care, and introduction of therapeutics have had surprisingly minimal effects on 60-day mortality prediction for hospitalized patients. Those predicted on hospital admission to do poorly demonstrate a rapid progression of illness, with transition to the ICU within the first 24 hours. Finally, although hospital mortality is the commonly quoted outcome metric for hospitalized patients, it underestimates the lethality of COVID-19, which continues to at least 60 days after hospital admission.

### Findings in Context

Crude mortality of hospitalized patients decreased throughout the pandemic, but once simple characteristics of patients being admitted were taken into account, predicted mortality was unchanged from the beginning of the pandemic to this time, suggesting that it is the profile of hospitalized patients that has changed and created a Simpson’s Paradox.^17^ We clearly demonstrate the continuous change of the at-risk population and have defined 7 risk groups that remained evident throughout the pandemic, although their constituents have fluctuated in number across the pandemic. These categories predict not only mortality but the escalation of care once patients are hospitalized.

It is becoming clear that despite emergence of SARS-CoV-2 variants and declining effectiveness of the initial vaccine platforms to generate vaccine-induced immunity, protection against hospital admission, and consequently COVID-related death, has remained surprisingly robust during evolution of the illness.^18^ Prevention of severe illness requiring hospitalization in the vaccinated cohort, as well as presumed slowly developing herd immunity in the general population and early treatment and mitigation of infection,^19^ are presumably censoring many otherwise high-risk patients from the cohort we are studying and contributing to the Simpson’s Paradox that is observed. Unfortunately, it is these same high-risk individuals with the greatest observed waning protection afforded by vaccination^20^ who still serve as the substrate for our study and continue to be identified by our model.

The at-admission risk model we generated uses simple, standard demographic data, medical history, vaccination status, symptoms, and routine admission laboratory data, yet impressively predicts outcome of hospitalized patients. Severity of subsequent disease can be fairly crudely estimated by symptom-complex (such as respiratory rate, oxygen saturation, blood pressure, heart rate, temperature, alertness), limited laboratory analysis (blood urea nitrogen, C-reactive protein), and demographics (age, sex), as several other models have demonstrated.^21–23^

Some of these models require a 24-hour period of assessment to become informed, which compromises their utility. Of these variables, routine hematologic and serum chemistry data are critical. During the “incubation” period, and perhaps in the early phase of the disease, hematologic constituents seem relatively unperturbed.^24^ In patients whose disease progresses to hospitalization, there is a profound derangement of serum chemistries and increase of inflammatory mediators and cytokines that give rise to a severe inflammatory response syndrome.^25, 26^

The genesis of life-threatening illness seems to be the induction of multiple interleukins, macrophage proinflammatory proteins and tumor necrosis factor-alpha, thrombosis and endothelial dysfunction, and platelet activation among innumerable other identified homeostatic derangements.^27^ At some transition point, which we suspect is just prior to hospital admission, individuals begin to declare their fate, as predicted by at-admission demographics, comorbidity, and laboratory values. Lymphopenia and neutrophilia are important predictors of severe disease, with decreased lymphocyte count at admission being associated with ICU destination, and higher neutrophil count identified as a risk factor for death.^28–30^ Moreover, increased levels of inflammatory markers at admission, such as C-reactive protein, procalcitonin, lactate dehydrogenase, and ferritin, seem to portend a more ominous prognosis.^24, 28, 31, 32^ Many of these variables have been similarly shown to predict poor outcome in our study and serve as reliable biomarkers of mortality to be used in risk calculators.

Although it is comforting that fewer patients are progressing to life-threatening illness, it remains concerning that mortality of a patient admitted in 3/2020 and predicted to be at high risk is essentially no different from that predicted for the same patient tomorrow. We are surprised by what we have found, given the types of resources we can marshal and the clinical strength of our hospital system. We can only speculate that we are starting late in the race to combat this disease. We are seeing many patients coming to the hospital already in extremis—dyspneic, lethargic, or dehydrated—and receiving rapidly escalating care soon after their arrival, suggesting severe disease at admission. It has been suggested that although viremia peaks within the first week of infection, peak host primary immunologic response may not be evident for perhaps as long as a week later.^33^

Appreciating that most patients will seek emergency room care only when the intensity of disease has progressed to severe illness, it could be anticipated that interventions targeting reducing viral load may not be effective if the viremia is no longer driving the disease process.^34, 35^ Recommendations for convalescent plasma administration for COVID-19 focus on early use^36^ and are consistent with early intervention being more favorable than later for similar therapeutics. Consequently, it is not surprising that anti-inflammatories such as corticosteroids, targeted toward the disease component presumably driving hospital outcome, have become a central and critically important intervention for hospitalized patients.^27, 28^

We postulate that the late risk of mortality after discharge is related to the initial COVID-19 disease and represents persisting disease and debility post-discharge or transfer as opposed to post-acute COVID-19 sequelae.^37^ This elevated risk above that expected in the U.S. population even beyond 60 days^38^ suggests the chronic and insidious nature of late COVID-19 mortality and may be part of post–COVID-19 conditions.^39, 40^

Finally, it is interesting that vaccination status did not protect patients from mortality once hospitalized. We are confident that vaccination has reduced hospital readmissions for COVID-19 and favorably contributes to the decreasing trend of severely ill patients. It has been speculated that host immune evasion in vaccinated patients is promoted by viral mutations occurring outside the spike coding region and that this is one mechanism of the apparent dissociation of vaccination status and outcome in those culled by hospital admission who populate this study.^41^ Unfortunately, vaccination as a variable is extraordinarily confounded by its timing, number of boosters received, and vaccine platform. None of this information was captured in our data set. This limits the appreciation of the contribution of vaccinations as the pandemic evolved.

### Limitations

This is a study of the COVID-19 experience in a single hospital system and is not necessarily generalizable nationwide. It focuses on patients hospitalized with severe or life-threatening COVID-19. Granular information, however, about the decision to admit a given patient is not available. Data elements recorded in the COVID-19 Registry reflect health-system decisions made at the beginning of the pandemic and were not modified to include potentially important variables such as certain presentation variables and laboratory tests such as ferritin, lactate dehydrogenase, coagulation factors, and patient-specific SARS-CoV-2 variant information. Because of lags in data entry into the COVID-19 Registry, risk factors currently identified as important in this study are not available in real time for clinical decision-making.

We restricted our analyses to the earliest available clinical information to enable generation of useful models that could predict mortality and direct allocation of resources. Thus, we did not consider additional hospital events, such as mechanical ventilation or extracorporeal membrane oxygenation. The study was conducted in real time during the pandemic, amid changing variants,^42^ changing thoughts about respiratory support, experimentation with therapeutics, and a variety of clinical trials. Consequently, care of patients across the pandemic has been difficult to standardize.

### Conclusions and Relevance

The evolution of the pandemic as addressed by us and others^23^ strongly suggests that COVID-19 remains lethal, albeit for a declining at-risk group of the general population. Observed mortality of patients hospitalized with COVID-19 appears to be decreasing over time, corroborating this. That there appear to be fewer patients who manifest severe disease in response to exposure and infection suggests that something has changed to favorably affect disease outcome. Clearly, dissemination of vaccination programs and herd immunity are at play, and there may be a change in viral virulence with development of variants that has led to a smaller fraction of the general population presenting with advanced disease. Early use of antiviral agents may sufficiently blunt the disease, and fewer people now need to seek advanced medical attention.

It is unlikely that we are done with COVID-19. Unfortunately, there is likely an underreporting of COVID-19–related mortality when only in-hospital outcomes are examined, as we find that risk of death persists well after hospital discharge. With continued emergence of variants having possibly different virulence, interval outcomes assessment is critical. The surprisingly nimble adaptability of the virus, and seemingly unchanging fate of those who develop severe hospital-requiring COVID-19, mandate that continued individual and public health efforts remain in place.

## Data Availability

Data used for this study include human research participant data that are sensitive and cannot be publicly shared due to legal and ethical restrictions by the Cleveland Clinic regulatory bodies, including the Institutional Review Board and legal counsel. In particular, variables such as date of testing or dates of hospitalization are HIPAA-protected health information and legally cannot be publicly shared. We will make our data sets available on request, under appropriate data use agreements with the specific parties interested in academic collaboration. Requests for data access can be made to Dr. Sudish C. Murthy.

## ACKNOWLEDGMENTS

The authors thank Alex Milinovich and Greg Strnad for management of the COVID-19 Registry, Ben Kramer for data verification, Beth Lieber for statistical programming, Leslie Jelinski for reference management, Brian Kohlbacher for graphic design expertise, and Tess Parry for editorial assistance.

## Appendix 1 Variables considered in analyses, with short names in parentheses, as seen on variable importance graphs (Figures 8A and 8C)

### Pre-COVID-19 Patient Characteristics

#### Demographics

Age (y) (age), sex (female), race (Black, White, Asian, mixed/other) (race_grp), Hispanic ethnicity (ethnicity), body mass index (kgom^-2^) (bmi)

#### Comorbidities

Smoking history (hx_smoke), pack-years of smoking (packyrs), chronic obstructive pulmonary disease, emphysema (copd_emp), chronic asthma (asthma), heart failure (hrtfail), coronary artery disease (cad), hypertension (htn), diabetes (diabetes), cancer (cancer), musculoskeletal disease (msclero), prior transplant (tx_hx), connective tissue disease (contxdz), inflammatory bowel disease (ibd), epilepsy (epilepsy), immune compromised (immu_dz), pregnant (pregnant)

#### Treatments and Medications

Has had a seasonal influenza vaccination (flu_shot), has had a pneumococcal pneumonia immunization (pneushot), pre-COVID-19 medications (nonsteroidal anti-inflammatory drug (nsaids), steroid (steroids), beta-blocker for hypertension (carvedil), angiotension-converting enzyme inhibitor (aceinhib), angiotensin receptor blocker (arb), selective serotonin reuptake inhibitor (paroxeti), melatonin (melatoni), colchicine (colchici)

### Hospital Admission Variables

Date of hospital admission (as count of days since 3/1/2020) (iv_daysa)

#### SARS-CoV-2 Variant

Pre-4/1/21: Alpha B.1.1.7; 4/1/2021 to 7/1/21: Alpha Q; 7/1/21 to 12/1/21: Delta B.1.617.2 and AY; 1/1/21 to 2/1/22: Omicron BA.1; 2/1/22 to 3/21/22: BA.1.1; 3/21/22 to 5/21/22: Omicron BA.2; 5/21/22 to 6/15/22: BA2.12.1

##### Symptoms and Laboratory Values at Hospital Admission

Symptoms (cough (cough), fatigue (fatigue), dyspnea (sob), diarrhea (diarrhea), loss of appetite (lossappt), vomiting (vomit), fever (fever01), sputum production (sputprod), flu-like symptoms (flu_symp), C-reactive protein (mgoL^-1^) (ad_crp), white blood cell count (10^9^•L^-1^) (ad_wbc), neutrophil count (10^9^•L^-1^) (ad_neutr), lymphocyte count (10^9^•L^-1^) (ad_lymoh), red blood cell count (10^12^•L^-1^) (ad_rbc), red blood cell distribution width (%) (ad_rdw), platelet count (10^9^•L^-1^) (ad_plate), hemoglobin (godL^-1^) (ad_hemog), hematocrit (%) (ad_hct), creatinine (mgodL^-1^) (ad_creat), blood urea nitrogen (mgodL^-1^) (ad_bun), albumin (godL^-1^) (ad_album), bilirubin (mgodL^-1^) (ad_bilir), alanine aminotransferase (IU•L^-1^) (ad_alt), aspartate aminotransferase (IU•L^-1^) (ad_ast), alkaline phosphatase (IU•L^-1^) (ad_alkph), potassium (mmol•L^-1^) (ad_potas), chloride (mEq•L^-1^) (ad_chlor), d-dimer (ngomL^-1^) (ad_ddime)

## Appendix 2 Alphabetic list of short names of variables considered in multivariable analyses and plots of variable importance

Appendix 2: Alphabetic list of short names of variables considered in multivariable analyses and plots of variable importance
|  |  |
| --- | --- |
| aceinhib | Angiotension-converting enzyme inhibitor |
| ad_album | Albumin ($\text{g}\cdot\text{dL}^{-1}$) |
| ad_alkph | Alkaline phosphatase ($\text{IU}\cdot\text{L}^{-1}$) |
| ad_alt | Alanine aminotransferase ($\text{IU}\cdot\text{L}^{-1}$) |
| ad_ast | Aspartate aminotransferase ($\text{IU}\cdot\text{L}^{-1}$) |
| ad_bilir | Bilirubin ($\text{mg}\cdot\text{dL}^{-1}$) |
| ad_bun | Blood urea nitrogen ($\text{mg}\cdot\text{dL}^{-1}$) |
| ad_chlor | Chloride ($\text{mEq}\cdot\text{L}^{-1}$) |
| ad_creat | Creatinine ($\text{mg}\cdot\text{dL}^{-1}$) |
| ad_crp | C-reactive protein ($\text{mg}\cdot\text{L}^{-1}$) |
| ad_ddime | D-dimer ($\text{ng}\cdot\text{mL}^{-1}$) |
| ad_hemog | Hemoglobin ($\text{g}\cdot\text{dL}^{-1}$) |
| ad_hct | Hematocrit (%) |
| ad_lymoh | Lymphocyte count ($10^9\cdot\text{L}^{-1}$) |
| ad_neutr | Neutrophil count ($10^9\cdot\text{L}^{-1}$) |
| ad_plate | Platelet count ($10^9\cdot\text{L}^{-1}$) |
| ad_potas | Potassium ($\text{mmol}\cdot\text{L}^{-1}$) |
| ad_rbc | Red blood cell count ($10^{12}\cdot\text{L}^{-1}$) |
| ad_rdw | Red blood cell distribution width (%) |
| ad_wbc | White blood cell count ($10^9\cdot\text{L}^{-1}$) |
| age | Age (y) |
| arb | Angiotensin receptor blocker |
| asthma | Chronic asthma |
| bmi | Body mass index ($\text{kg}\cdot\text{m}^{-2}$) |
| cad | Coronary artery disease |
| cancer | Cancer |
| carvedil | Beta-blocker for hypertension |
| colchici | Colchicine |
| contxdz | Connective tissue disease |
| copd_emp | Chronic obstructive pulmonary disease, emphysema |
| cough | Cough |
| diabetes | Diabetes |
| diarrhea | Diarrhea |
| epilepsy | Epilepsy |
| ethnicity | Hispanic ethnicity |
| fatigue | Fatigue |
| female | Sex |
| fever01 | Fever |
| flu_shot | Has had a seasonal influenza vaccination |
| flu_symp | Flu-like symptoms |
| hrtfail | Heart failure |
| htn | Hypertension |
| hx_smoke | Smoking history |
| ibd | Inflammatory bowel disease |
| immu_dz | Immune compromised |
| iv_daysa | Date of hospital admission (as count of days since 3/1/2020) |
| lossappt | Loss of appetite |
| melatoni | Melatonin |
| msclero | Musculoskeletal disease |
| nsaids | Current medications: nonsteroidal anti-inflammatory drug (nsaids) |
| packyrs | Pack-years of smoking |
| paroxeti | Selective serotonin reuptake inhibitor |
| pneushot | Has had a pneumococcal pneumonia immunization |
| pregnant | Pregnant |
| race_grp | Race (Black, White, Asian, mixed/other) |
| steroids | Current medications: steroid |
| sob | Dyspnea |
| sputprod | Sputum production |
| tx_hx | Prior transplant |
| variant | Pre-4/1/21: Alpha B.1.1.7<br>4/1/2021 to 7/1/21: Alpha Q<br>7/1/21 to 12/1.21: Delta B.1.617.2 and AY<br>12/1/21 to 2/1/22: Omicron BA.1<br>2/1/22 to 3/21/22: BA.1.1<br>3/21/22 to 5/21/22: Omicron BA.2<br>5/21/22 to 6/1/22: BA2.12.1 |
| vomit | Vomiting |

