## Supplemental material for "Evolution of Life-Threatening COVID-19 as the SARS-CoV-2 Pandemic Has Progressed"

**Table E1:** In-hospital events, medications, and discharge location of persons admitted to Cleveland Clinic Health System hospitals with COVID-19

**Figure E1:** Completeness of follow-up. Red x's represent deaths and blue circles living patients positioned at time of last follow-up, with a closing date of 7/1/2022. **A)** Patients treated in Ohio. **B)** Addition of the 2,216 Florida patients, with their last cross-sectional follow-up as of 1/1/2022. **C)** Patients treated in Ohio, zoomed in at 60 days after hospital admission. **D)** Addition of patients treated in Florida, zoomed in at 60 days after hospital admission.

**Figure E2:** Partial dependency plots of the relationship of predicted 60-day risk-adjusted mortality and age at hospital admission for COVID-19 based on the pre–COVID-19 mortality model incorporating demographics, medical history, medications, and vaccination status (red symbols and line), and the at-admission mortality model, also incorporating symptoms and results of routine laboratory tests at hospital admission, reflecting patients' physiologic response to SARS-CoV-2 (blue symbols and line). Note blunting of the age relationship in the at-admission model vs. pre–COVID-19 model when patients' response to the virus is taken into account.

**Table E1: In-hospital events, medications, and discharge location of persons admitted to Cleveland Clinic Health System hospitals with COVID-19**

| | COVID-19 Admitted (N=13,327) | |
|---|---|---|
| Variable | n[a] | No. (%) |
| ***In-hospital events*** | | |
| High-flow oxygen | 26,872 | 4,307 (16) |
| Mechanical ventilation | 26,872 | 2,555 (9.5) |
| Admission to intensive care unit | 26,872 | 6,081 (23) |
| Extracorporeal membrane oxygenation | 26,872 | 13 (0.048) |
| ***Medications*** | | |
| Steroids | 26,872 | 15,821 (59) |
| Vitamin D | 26,872 | 4,034 (15) |
| Zinc | 26,872 | 907 (3.4) |
| Hydroxychloroquine | 26,872 | 637 (2.4) |
| Tocilizumab | 26,872 | 452 (1.7) |
| Remdesivir | 26,872 | 211 (0.79) |
| Lopinavir/Ritonavir | 26,872 | 9 (0.033) |
| Oral ribavirin | 26,872 | 1 (0.0037) |
| ***Discharge Location*** | | |
| Home | 26,164 | 16,631 (63) |
| Skilled nursing facility | 26,164 | 3,489 (13) |
| Other facility | 26,164 | 962 (3.7) |
| Died in hospital | 26,164 | 1,503 (5.7) |

a. Patients with data available.

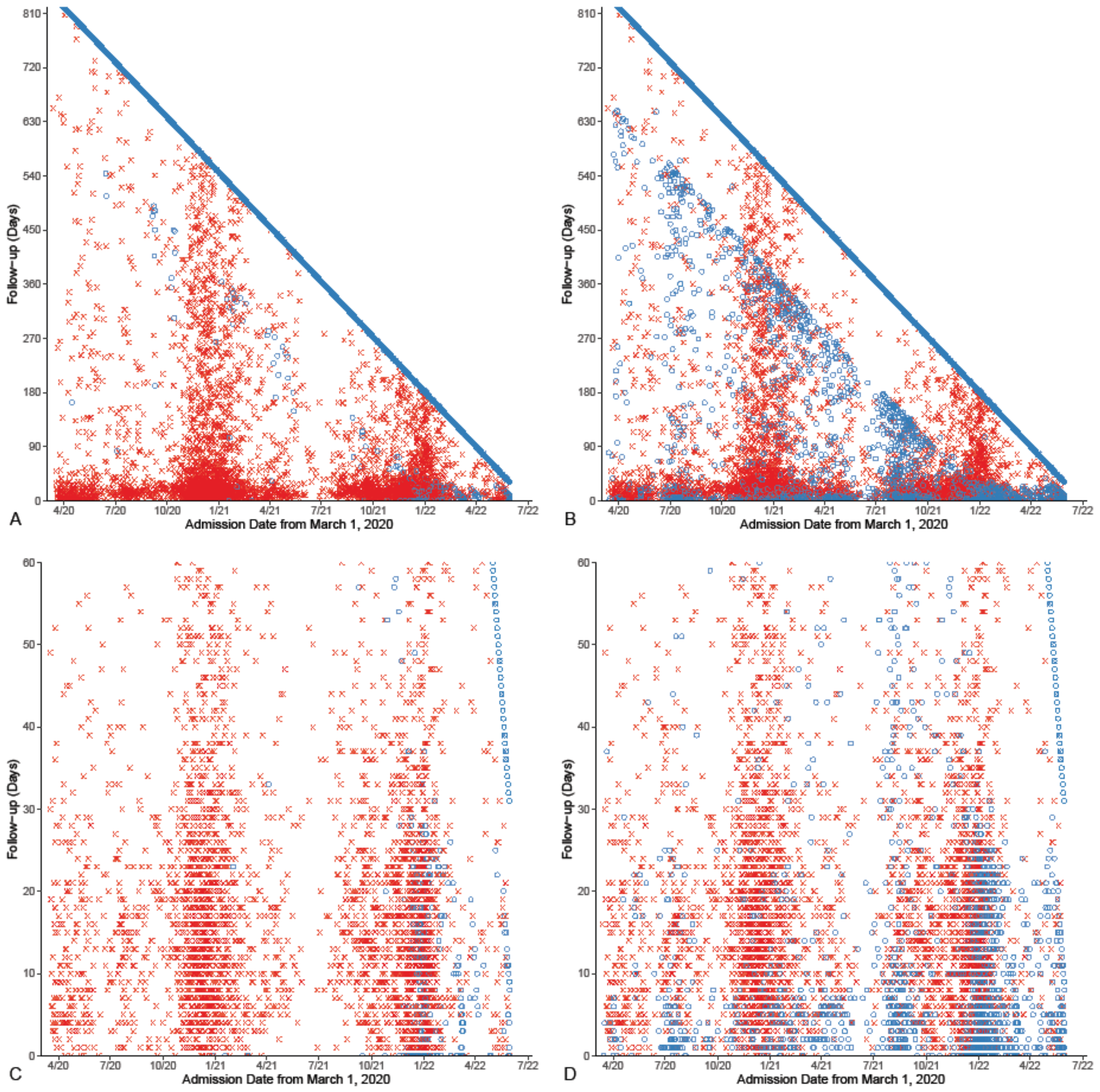


**Figure E1:** Completeness of follow-up. Red x's represent deaths and blue circles living patients positioned at time of last follow-up, with a closing date of 7/1/2022. **A)** Patients treated in Ohio. **B)** Addition of the 2,216 Florida patients, with their last cross-sectional follow-up as of 1/1/2022. **C)** Patients treated in Ohio, zoomed in at 60-days post-hospital admission. **D)** Addition of patients treated in Florida, zoomed in at 60-days post-hospital admission.

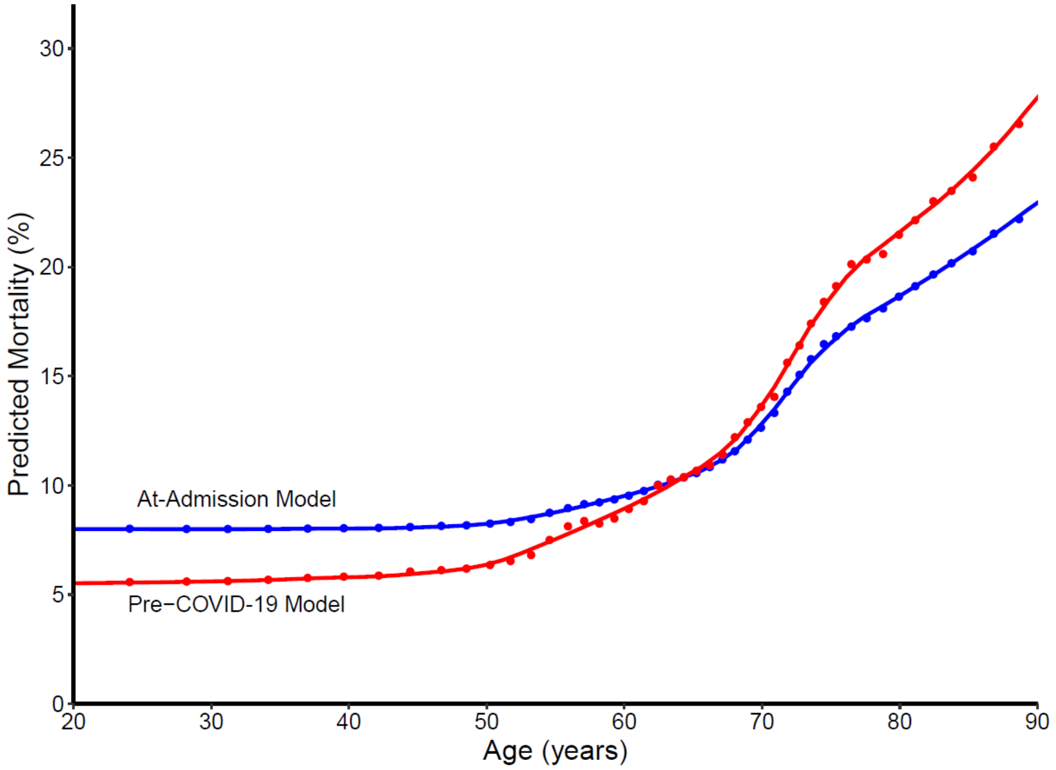


**Figure E2:** Partial dependency plots of the relationship of predicted 60-day risk-adjusted mortality and age at hospital admission for COVID-19 based on the pre–COVID-19 mortality model incorporating demographics, medical history, medications, and vaccination status (red symbols and line), and the at-admission mortality model, also incorporating symptoms and results of routine laboratory tests at hospital admission, reflecting patients' physiologic response to SARS-CoV-2 (blue symbols and line). Note blunting of the age relationship in the at-admission model vs. pre–COVID-19 model when patients' response to the virus is taken into account.
